# Patterns and functional consequences of antibody speciation in maternal-fetal transfer of coronavirus-specific humoral immunity

**DOI:** 10.1101/2024.09.12.24313591

**Authors:** Andrew P. Hederman, Hannah M. Brookes, Harini Natarajan, Leo Heyndrickx, Kevin K. Ariën, Joshua A. Weiner, Amihai Rottenstreich, Gila Zarbiv, Dana Wolf, Tessa Goetghebuer, Arnaud Marchant, Margaret E. Ackerman

## Abstract

Maternal antibodies serve as a temporary form of inherited immunity, providing humoral protection to vulnerable neonates. Whereas IgG is actively transferred up a concentration gradient via the neonatal Fc Receptor (FcRn), maternal IgA and IgM are typically excluded from fetal circulation. Further, not all IgG molecules exhibit the same transfer efficiency, being influenced by subclass, Fab and Fc domain glycosylation, antigen-specificity, and the temporal dynamics of maternal antibody responses. Here, we investigate the phenotypes and functions of maternal and cord blood antibodies induced by SARS-CoV-2 infection and compare them to those induced by mRNA vaccination, focusing on breadth of antigen recognition and antiviral functions including neutralization and effector function. While cord blood coronavirus-specific antibody functional breadth and potency appeared to be more compromised than binding breadth and potency in both groups, vaccination induced substantially greater function and breadth in cord blood than did natural infection. These functional phenotypes were associated with speciation of the maternal serum repertoires, as some IgG subpopulations were enriched while others were relatively depleted. Relevant to the continued protection of vulnerable populations in the context of a diversifying pathogen, greater breadth was observed for antibody effector functions than for neutralization, and these activities were associated with greater affinity for antigen. This work provides insights into the functional breadth of maternal-fetal antibody responses in the context of novel mRNA vaccines and a recently emerged pathogen that is likely to be a public health burden for the foreseeable future.

## Introduction

The current COVID-19 pandemic continues to infect and cause severe disease in many individuals. While the global death rate has declined, it is estimated that even in a relatively well-vaccinated population like the United States, 3,000 deaths per month are attributable to Severe Acute Respiratory Syndrome coronavirus-2 (SARS-CoV-2), the virus responsible for COVID-19^1^. The virus’ sequence has diverged over time into numerous variants with differing degrees of susceptibility to antibodies induced by exposure to prior variants. Notable variants, including Alpha (B.1.1.7), Beta (B.1.351), Gamma (P.1), Delta (1.617.2), and Omicron (B.1.1.529) have shown the ability, albeit at varying levels, to evade neutralizing antibody responses^2–11^. Across each of these waves of prevalent viral variants, certain populations of individuals remain at greater risk than others for severe COVID-19 following infection. Fortunately, while initial vaccine clinical trials did not include pregnant participants, recent data has shown that following vaccination, pregnant women achieve robust antibody responses that are comparable to non-pregnant individuals, while maintaining similar safety profiles, and that vaccination is highly effective in preventing severe disease and death in birthing women^12–22^.

Further, because mammals have evolved to transmit maternal humoral immunity to the fetus/neonate in the form of antibodies transferred across the placenta and via breastmilk, maternal vaccination has the multifaceted goal of not only protecting the mother but also of generating a sufficiently robust antibody response to protect the fetus in utero as well as the neonate during early life^23^. Indeed, whether induced by infection or vaccination, the “inheritance” of maternal antibody is a crucial component of neonatal immunity early in life against infectious diseases^24,25^. While not especially susceptible to COVID-19 morbidity and mortality, neonates and children remain at risk of long term post-acute sequelae of COVID-19 (PASC)^26^. Although rare, these complications can be severe including multisystem inflammatory syndrome in children (MIS-C)^27^. Additionally, while both maternal infection and maternal vaccination are known to influence maternal and neonatal infection risk, at least for some variants^28–30^, they induce maternal antibody responses with a number of differing attributes, providing an opportunity to evaluate transfer biases and their impacts.

Transport of IgG across the placenta is mediated by binding to the neonatal Fc receptor (FcRn) expressed on syncytiotrophoblast cells^31–34^. Intriguingly, these antibodies do not bind to FcRn on the cell surface, but rather are rescued from degradation following fluid phase uptake, internalization, and acidification of endosomal compartments by a pH-dependent protonation of the Fc domain, which results in increased affinity to FcRn^35^. As FcRn is then sorted and cycled back to the cell surface, it salvages bound IgG, releasing it at the neutral pH of the extracellular environment. The efficiency of this rescue and transfer across the otherwise highly selective barrier between generations is associated with many factors, including maternal IgG levels over time, IgG subclass and allotype, post-translational modifications on both antigen-binding and Fc domains, and antigen-specificity^36–43^. While some of these factors are thought to drive these differences by directly altering FcRn binding affinity or kinetics, the mechanism(s) whereby other factors relate to transfer phenotypes remains incompletely understood^44–46^. Studies have started to look at the transfer of antibodies following maternal mRNA vaccination against SARS-CoV-2^38,47–49^; however, a more thorough analysis of the phenotypes and activity of the transferred antibodies could provide insight into both protection of neonates from COVID-19 as well as to other infectious diseases by informing on attributes and activities of maternal immune responses that might provide for the best defense of vulnerable neonates.

Previous analysis of vaccinated pregnant women has revealed generation of neutralizing SARS-CoV-2 specific antibodies as well as antibodies capable of eliciting Fc-mediated effector functions in mothers^50–52^. These same antibody attributes are typically present in cord blood at delivery^51^. In other contexts, however, they are not always present at the levels expected based on antibody titers, indicating that while generally well-correlated with maternal serum antibodies at the time of delivery, relative activity of antibodies in cord blood are distinct. Studies reporting differences in quantity and quality of transferred antibodies have cited numerous factors including maternal antibody levels, FcRn expression level, IgG glycosylation, IgG subclass, and antigen-specificity, among others, as potential explanations^36–43^. While it is well understood that vaccination-and natural infection-induced antibody responses differ in terms of mucosal compartmentalization, induced isotype balances, and epitope-specificities, what remains less well studied is the breadth of the antibody functional response in mothers as it compares to matched cord blood to SARS-CoV-2 variants of concern (VOC) following these exposures^53–58^. These factors are important as the virus continues to diversify, population-level uptake of updated and booster vaccinations decline^59^, and efforts continue toward a universal vaccine^60^. Overall, understanding the breadth of the antibody functional response in vaccinated and convalescent mothers with corresponding matched cord blood samples can provide new insights into how neonates may benefit from maternal antibodies, in the context of compromised or lost neutralization activity associated with viral variation over time and the absence of pathogen-specific IgA and IgM. These insights into the maternal and inherited antibody repertoires have implications for how vaccines can be most effectively developed as new variants continue to emerge for SARS-CoV-2 as well as for other infectious agents posing risks to neonates and infants.

## Results

### Distinct antibody responses among cord blood samples from vaccinated and convalescent mothers to SARS-CoV-2 variants

To explore antibody profiles to SARS-CoV-2 variants, maternal and matched cord blood serum samples were collected from vaccinated (n=50, Hadassah Medical Center, Israel) and convalescent maternal study participants (n=38, CHU St. Pierre, Belgium) after immunization against or infection by SARS-CoV-2 in the third trimester early in the pandemic (**Supplemental Table 1**). These samples were profiled for antibody magnitude, specificity, and Fc domain characteristics of SARS-CoV-specific IgM, IgA, and IgG across a panel of variants and pertussis and tetanus toxoids as control or comparator specificities that are also relevant to neonatal health (**Supplemental Table 2**). We first explored the antibody binding responses to SARS-CoV-2 Wuhan and VOC in maternal and cord blood as a representation of antibody profiles in neonates born to vaccinated and convalescent mothers. Antibody profiles differed in association with both maternal exposure history and sample type, as shown from the distinct clustering by Uniform Manifold Approximation (UMAP) (**Figure 1A**). Responses among maternal blood samples clustered closely together, while those in cord samples formed a distinct cluster. Within each of these sample type groups, vaccinated and convalescent subjects formed distinct sub-clusters.

**Figure 1:**
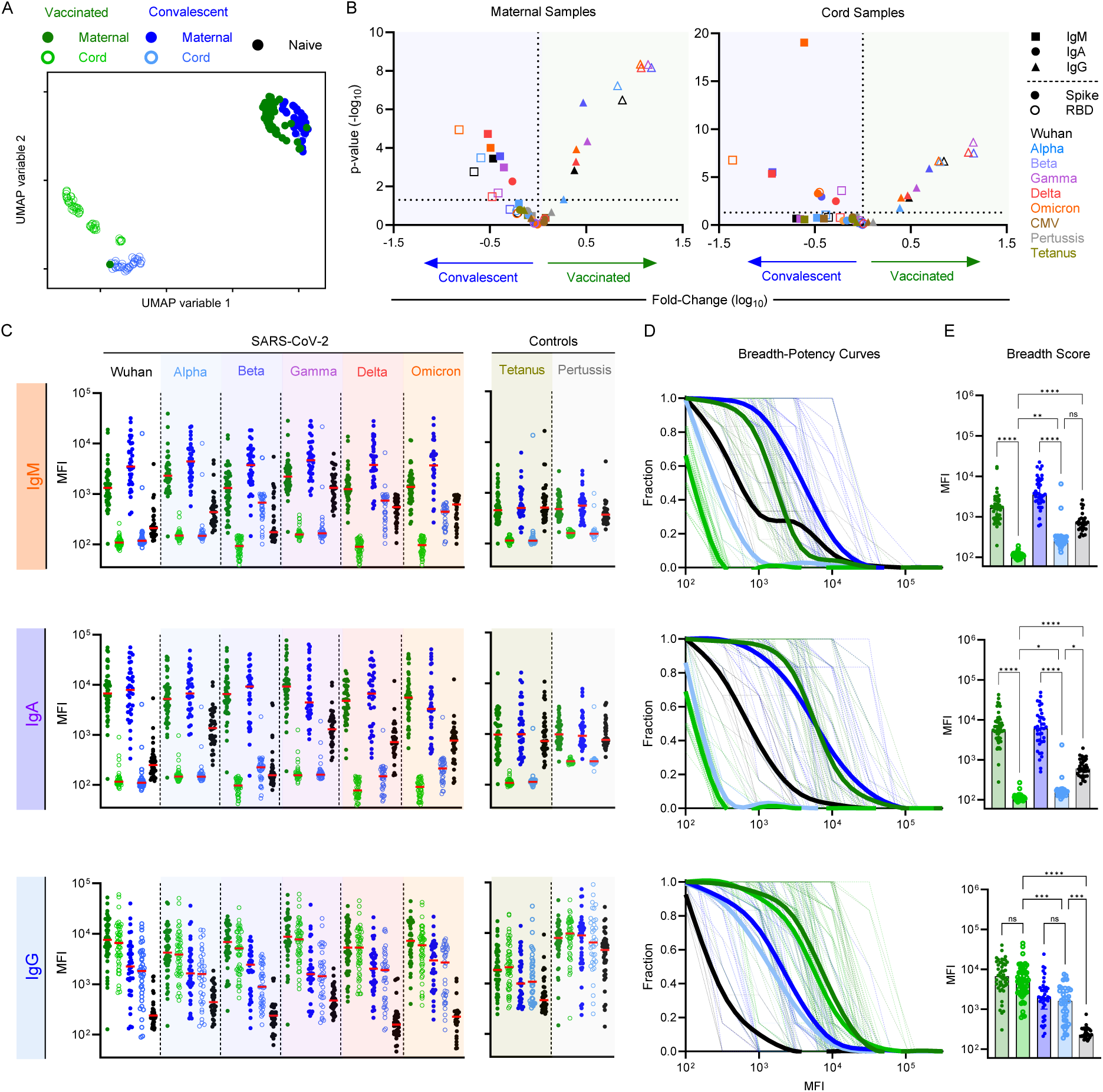
Antibody isotype, magnitude, and breadth across SARS-CoV-2 VOC in maternal and cord blood samples following vaccination or natural infection. **A.** Coronavirus-specific antibody response features after dimensional reduction in maternal (filled) and cord (open) samples among convalescent (n = 38) (blue) or vaccinated (n = 50) (green) individuals. **B.** Volcano plot presenting the fold-change (x-axis) and statistical significance (Mann Whitney test, y-axis) of differences between convalescent and vaccinated participants. Antibody isotype is indicated by shape, with RBD and whole spike indicated in hollow and filled symbols, respectively. SARS-CoV-2 variant is indicated by color. **C.** Median Fluorescent Intensity (MFI) of IgG responses to spike of SARS CoV-2 VOCs as defined by multiplex assay for IgM (top), IgA (middle), and IgG (bottom). Responses among SARS-CoV-2 naïve subjects (n = 38) are shown in black. Bar indicates the median response. **D.** Breadth-potency curves represent the fraction of subjects with a response exceeding a given level for IgG antibody responses across the panel of VOC. Population means are shown with a thick line, and individual subjects illustrated in thin lines. **E.** IgG breadth scores for each subject. Bar indicates the median. Statistical significance was defined by ANOVA Kruskal–Wallis test with Dunn’s correction and α=0.05 (*p<0.05, **p<0.01, ***p<0.001, ****p<0.0001).

Among individual features of the antibody response that were assessed, higher levels of variant-specific IgM but lower levels of variant-specific IgG antibodies were observed in maternal blood from convalescent as compared to vaccinated subjects (**Figure 1B**). Intriguingly, despite the expectation that IgA and IgM would be absent from cord blood samples, some of these responses were nonetheless also elevated in cord blood samples from neonates whose mothers were convalescent (**Figure 1B**). Given these surprising results, responses measured in each sample were compared with blood samples from naïve subjects across each isotype for SARS-CoV-2 variants and control antigens (**Figure 1C**). In general, IgM and IgA reactivity toward all of the antigens tested were considerably lower in cord blood than even serum samples from SARS-CoV-2 naïve subjects, consistent with the presence of natural or cross-reactive IgM and IgA antibodies with some ability to bind all of the antigens tested, and the expected lack of IgA and IgM in cord blood. However, levels were not the same in cord blood samples from vaccinated as compared to convalescent dyads for a specific subset of SARS-CoV-2 antigens, including beta, delta, and omicron variants. Elevated IgM and IgA responses to these three antigens in cord blood of convalescent mothers were sufficient to lead to greater activity in both breadth-potency curves (**Figure 1D**), as well as breadth scores (**Figure 1E**). Similar elevations were also observed for IgM binding to the receptor binding domain (RBD) for gamma, delta, and omicron and for IgA binding to omicron (**Supplemental Figure 1**). Neither of the control antigens tested showed this pattern, suggesting that this signal is not the result of transfer of small quantities of these isotypes stemming from maternal infection. An explanation for this phenotype was not readily apparent, though some combination of specific mutations, distinct mutations in specific positions, conformationally-distinct epitopes, post-translational modifications, or other factors may contribute to these unique binding profiles.

In contrast to IgM and IgA responses, the breadth of IgG responses was elevated in association with vaccination as compared to infection for maternal blood, as previously reported for this cohort. This elevation was also apparent in cord blood (**Figure 1D-E**). For both exposure histories, IgG binding breadth was comparable between maternal and cord blood (**Figure 1C-E**). The breadth of RBD-specific IgG also exhibited the same consistency between maternal and cord blood within each group **(Supplemental Figure 1**), further demonstrating the efficiency of placental transfer even when seroconversion does not occur until during the third trimester. In sum, greater coverage of diverse variants is expected in infants whose mothers were immunized rather than infected based on their elevated IgG responses.

### IgG subclass and FcγR breadth of antibodies in cord blood to SARS-CoV-2 variants differs between vaccination and natural infection

Given the stark differences in IgG binding breadth associated with exposure history, we explored the breadth of IgG subclasses and Fcγ Receptor (FcγR) binding of spike-(S), RBD-, and control antigen-specific antibodies in cord blood samples (**Figure 2**, **Supplemental Figures 2-3**). IgG1 and IgG3 responses exhibited the greatest breadth, followed by IgG2; IgG4 responses were uncommon (**Figure 2A**, **left; Supplemental Figure 2**). Comparing between exposure histories, while breadth scores for all subclasses were higher following maternal vaccination than infection, this difference was only statistically significant for IgG3 (**Figure 2B**, **top**). FcγR binding antibody breadth-potency curves were similar among the receptors tested (**Figure 2A**, **right**), and like IgG3, breadth scores were significantly increased in cord blood following maternal vaccination as compared to infection (**Figure 2B**, **bottom**). Given the varying roles of these subclasses and FcγR to antibody effector functions, these differences may be relevant for the *in vivo* antiviral activity of cord blood antibodies.

**Figure 2:**
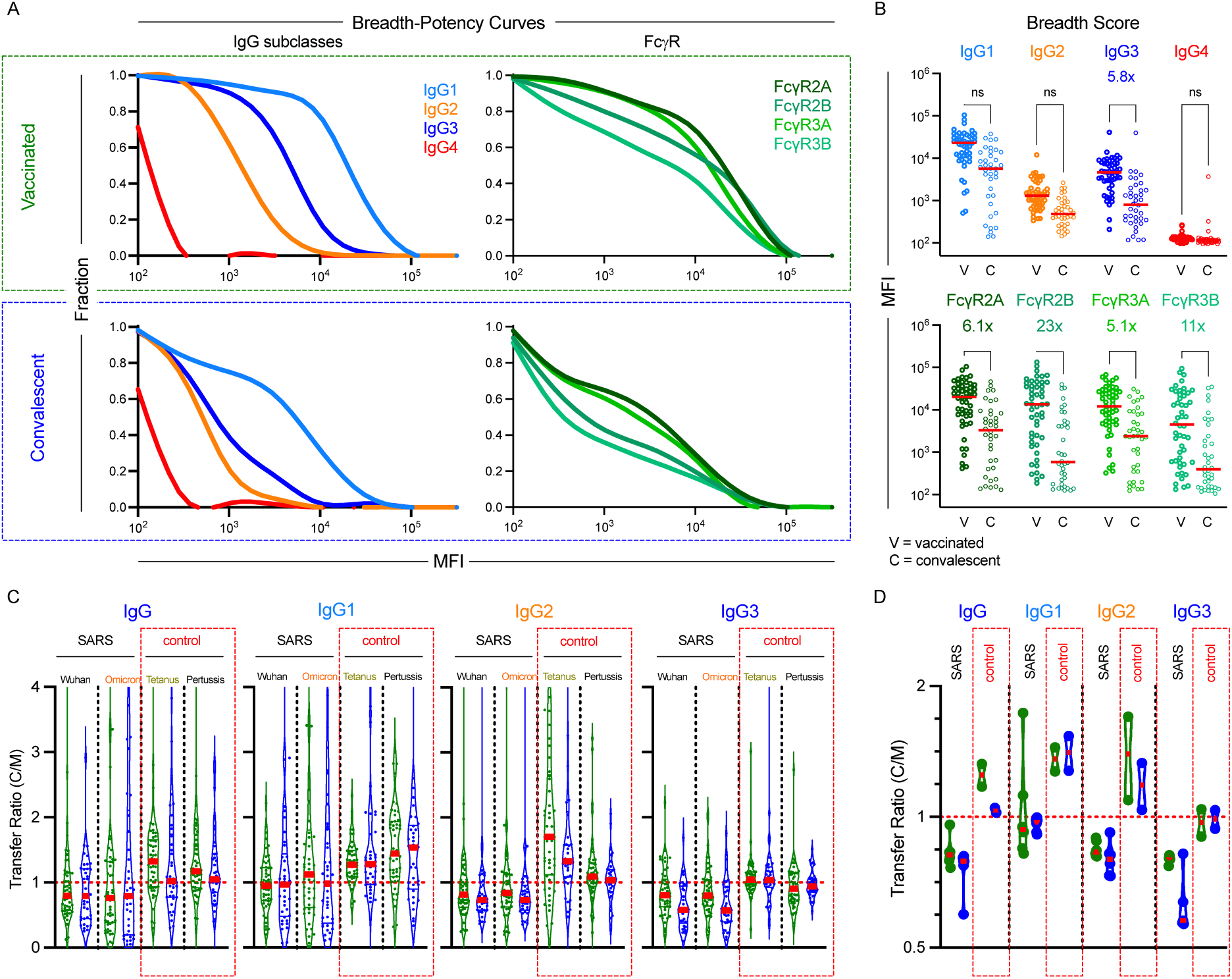
Vaccination and infection elicit distinct IgG subclass and Fc receptor breadth in cord blood. **A.** Breadth–potency curves representing the fraction of subjects with a response exceeding a given level for each IgG subclass (left) and for binding to FcR (right) across the panel of VOC in cord blood samples following maternal vaccination or infection. Population mean is shown with a thick line, and individual subjects are illustrated in thin lines. **B.** IgG subclass (top) and FcR binding (bottom) breadth scores for each cord blood samples following maternal vaccination (filled) or infection (hollow). Statistical significance was defined by ANOVA Kruskal–Wallis test with Dunn’s correction and α=0.05 (**p<0.01, ***p<0.001, ****p<0.0001). **C.** Transfer ratio (cord/maternal levels) of antigen-specific (SARS: Wuhan and Omicron Spike; Control: Pertussis and Tetanus) IgG subclasses in vaccinated (green) and convalescent (blue) dyads. **D**. Median transfer ratios of each SARS-CoV-2 spike VOC as compared to control antigens by subclass. Bars indicate median.

**Figure 3:**
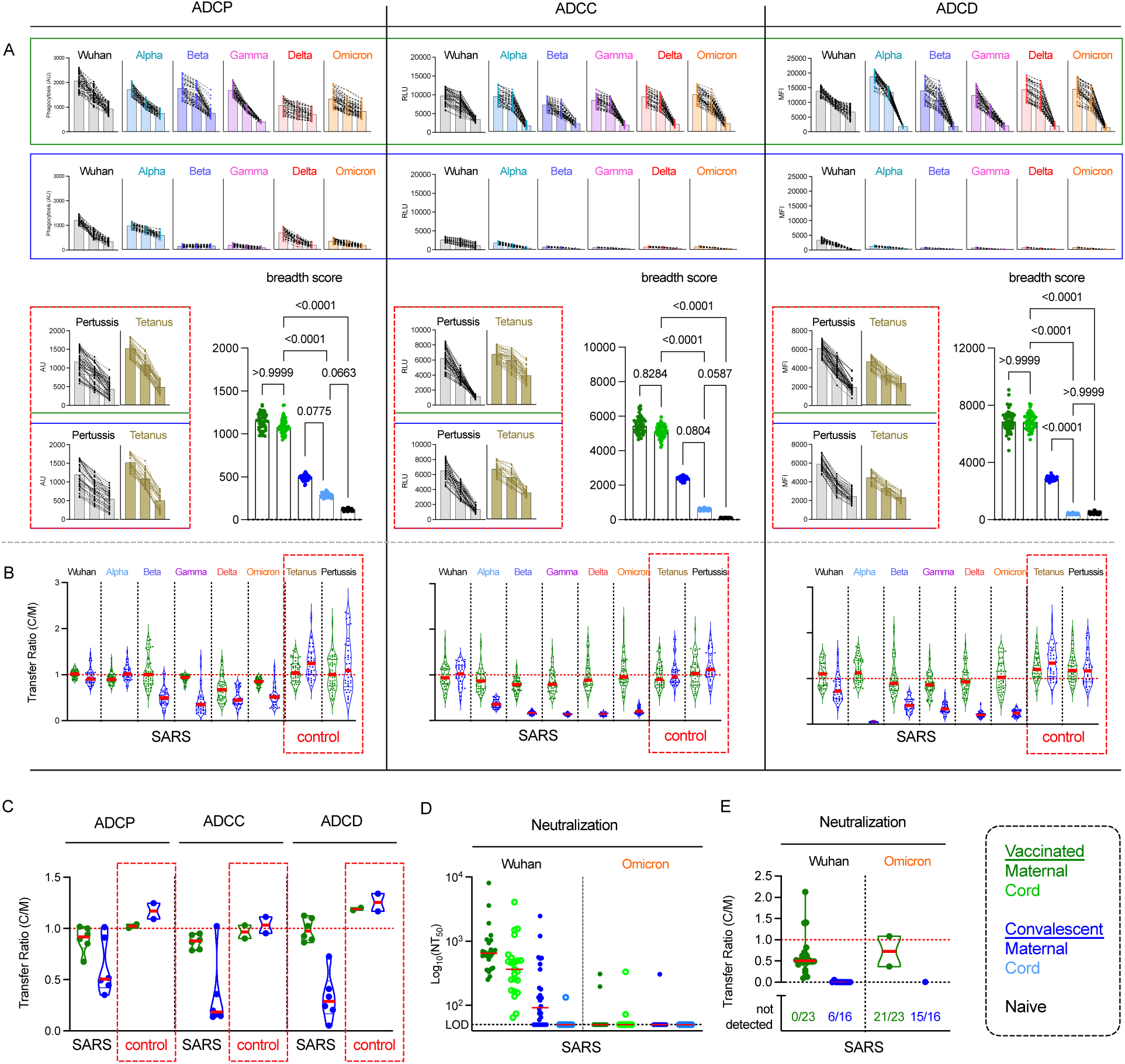
mRNA vaccination results in superior breadth of SARS-CoV-2-specific Ab effector function and neutralization in cord blood. **A.** Ab effector functions in cord blood from vaccinated (top, green shading) and convalescent (bottom, blue shading) cord blood for SARS spike variants or pertussis and tetanus control (red box) antigens. Phagocytosis, ADCC, and Complement deposition activities were assessed at each of three serum dilutions (1:50, 1:100, 1:250). Functional activity is reported in arbitrary units (AU), relative light units (RLU), and median fluorescent intensity (MFI). **Inset**. Functional breadth scores across variants in maternal, cord, and naïve subject samples. Statistical significance was defined by ANOVA Kruskal–Wallis test with Dunn’s correction and α = 0.05. **B-C.** Transfer ratios (cord/maternal) of Fc effector functions to indicated antigens in vaccinated (green) or convalescent (blue) dyads at the 1:50 dilution for individual antigens (**B**) and for the set of SARS and control antigens (**C**). **D.** Neutralization titers (NT_50_) observed for a subset of vaccinated (n=23) (green) and convalescent (n=26) (blue) maternal and cord samples against Wuhan (black) and Omicron (orange) strains. The limit of detection (LOD) is indicated by the horizontal dotted line. **E.** Transfer ratios of neutralization activity. Bars indicates median. Unless otherwise noted, data presented includes samples from 37 naive subjects, and 50 vaccinated and 38 convalescent dyads.

### Biases in the specificity and subclasses of well-transferred IgG

We next explored IgG transfer ratios (cord/maternal responses) to evaluate the extent to which antibody sub-populations were differentially transferred to cord blood when mothers were exposed to spike by natural infection or vaccination. Whereas total IgG specific for tetanus and pertussis control antigens was enriched in cord blood (**Figure 2C-D**), there was less total IgG specific for SARS-CoV-2 S protein present in cord blood than maternal blood for both Wuhan and the most distant VOC, Omicron (**Figure 2C**). Indeed, for dyads exposed to SARS by either vaccination or natural infection, median levels of total IgG binding to S were decreased in cord as compared to maternal blood (**Figure 2D**). Among the IgG subclasses, IgG1 was better transferred than IgG2, which was in turn better transferred than IgG3. This pattern or relative differences was consistent across specificities and groups (**Figure 2C-D**, **Supplemental Figure 4**). However, the absolute magnitudes differed: transfer efficiency of pertussis and tetanus-specific IgG1, IgG2, and IgG3 was greater than that of SARS-CoV-2-specific subclasses, and transfer efficiency in vaccinated dyads tended to be greater than that observed in convalescent dyads.

### mRNA vaccination shows increased antibody Fc effector functions and transfer of functional antibodies compared to natural infection

Given these biases in SARS-CoV-2 specific IgG phenotypes between maternal and cord blood samples, we next defined the ability of antibodies in cord blood from vaccinated and convalescent dyads to elicit Fc-mediated effector functions in *in vitro* assays. For each sample, we measured phagocytosis (ADCP), antibody dependent cellular cytotoxicity (ADCC), and complement deposition (ADCD) at three serum concentrations (**Figure 3A**, compared to maternal levels and for RBD in **Supplemental Figures 5-6**). For cord blood samples from vaccinated dyads, functional activity was well conserved across variants, whereas for the convalescent dyad cord blood samples, there was low activity against the Wuhan strain, and considerable reduction apparent in functional activity to most VOC across all effector functions. In contrast to SARS-CoV-2-specific responses, both vaccinated and convalescent cord blood samples showed robust functional activity to both tetanus and pertussis control antigens across all three functional assays (**Figure 3A**). Unlike SARS CoV-2-specificities, levels of activity were similar for the control antigens in cord blood drawn after maternal infection as after maternal vaccination.

In vaccinated dyads, functional breadth scores were similar between maternal and cord blood samples for both spike (**Figure 3A**) and RBD antigens (**Supplemental Figure 6**), suggesting that Fc mediated effector functions induced by vaccination and transferred *in utero* can be quite broad. However, like the IgG binding responses, functional breadth was greater in cord blood from vaccinated than convalescent dyads (**Figure 3A**, **inset**). For ADCP, ADCC, and ADCD, vaccinated maternal-cord pairs showed transfer ratios that tended be around one, with slightly higher values for closer VOC, and lower values for more distant VOC (**Figure 3B-C**, **Supplemental Figure 7**). Convalescent dyads showed a significantly different profile, with transfer ratios of around one for the Wuhan spike, but a significant reduction in transfer ratio of functional antibodies against VOC. Thus, the decreased magnitude of antibody responses in convalescent mothers is further compounded by decreased transfer efficiency, leaving neonates with considerably lower antibody effector activity against VOC. Again, in contrast to observations for SARS-CoV-2 antigens, control antigens typically exhibited functional transfer ratios greater than or equal to one in both groups (**Figure 3B-C**).

### Effector functions may contribute to protection of neonates in the absence of neutralization

Although loss of neutralization has been reported elsewhere for the Omicron variant^61^, this loss was confirmed for the participants in this study. Neutralization tests against the Wuhan and Omicron variants were performed for a subset of subjects (**Figure 3D**). Further, cord blood samples exhibited lower neutralization activity than did maternal blood against the Wuhan strain, consistent with prior reports^62^. Neutralization activity was greater for vaccinated as compared to convalescent dyads, and limited neutralization was observed against the Omicron variant. The transfer efficiency of neutralizing antibodies was poorer than that of antibodies with effector function; it was also lower in convalescent than vaccinated dyads (**Figure 3E**). Collectively, these results suggest that effector function has the potential to contribute to protection of neonates in cases where neutralization activity is lost or insufficient.

### Antibody functions are differentially mediated by immunoglobulin isotype

With prior data showing differences in immunoglobulin (Ig) isotype binding in cord as compared to maternal blood and functional and transfer efficiency differences associated with maternal exposure history, we next explored the role each isotype played in eliciting effector functions (**Figure 4**). Serum from maternal and cord blood was depleted of IgM, IgA, or IgG and tested for each effector function following confirmation of the efficiency (**Figure 4A**) and specificity (**Supplemental Figures 8-9**) of depletion. Functional responses for maternal and cord blood samples from vaccinated dyads were completely dependent on IgG, as depleted samples showed a near complete loss of functional activity (**Figure 4B-D**). In contrast, despite the presence of these additional isotypes, depletion of IgM or IgA from vaccinated dyads had no effect on these effector functions. Maternal and cord blood samples from convalescent dyads also showed dependence on IgG for ADCP and ADCC function (**Figure 4B-C**), but exhibited a unique profile for ADCD. For the maternal convalescent group, ADCD activity was significantly reduced by IgG depletion, but the magnitude of this reduction was relatively smaller than for other sample types, accounting for only about 10-25% of ADCD activity, the majority of which could instead be attributed to IgM (**Figure 4D**). Collectively, this data showed that ADCD activity was variably induced by IgM or IgG isotypes, depending on antigen exposure history.

**Figure 4:**
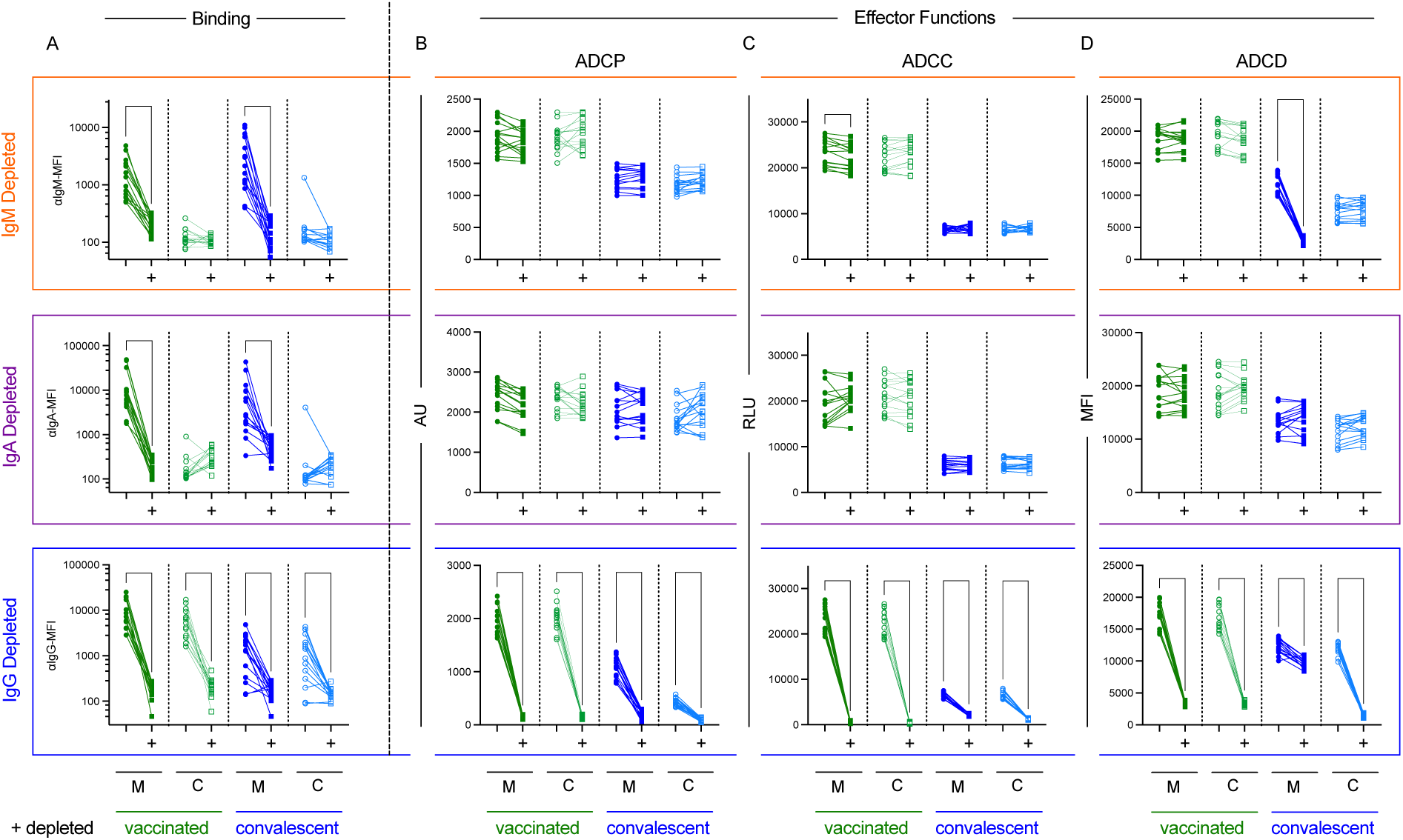
Contributions of each isotype to antibody effector functions in cord and maternal blood. Maternal (M) and cord (C) blood samples from vaccinated (green, n=15) and convalescent (n=15) dyads depleted of IgM (top), IgA (center), and IgG (bottom) antibodies. **A-D.** Binding levels (**A**, left) and effector functions (**B-D**) (right) to Wuhan spike protein were measured on mock and depleted (+) samples for each isotype to measure efficiency of each depletion and the impact on antibody activity against Wuhan spike antigen. Mock and depleted (+) samples were compared using a paired mixed effect model corrected for multiple hypothesis testing using the Benjamini, Krieger, and Yekutieli to control the false discovery rate (*q<Q, where Q=0.05). Functional activity is reported in arbitrary units (AU), relative light units (RLU), and median fluorescent intensity (MFI).

### Functional responses to endemic and emergent pathogenic coronaviruses

Lastly, induction of immunity beyond SARS-CoV-2 VOC, to other sarbeco-and further CoV families is clinically desirable. The breadth of recognition of SARS-CoV-1 and endemic CoV spike proteins are known to vary in association with SARS-CoV-2 spike exposure history^63,64^. The differences in antibody functional breadth among vaccinated and convalescent dyads across CoV-2 variants led us to explore to what extent this breadth encompasses more distant CoV. We tested binding and effector functions of antibody responses specific for endemic beta CoV HKU1, and OC43, and alpha CoV NL63, and 229E, along with emergent pathogenic SARS-CoV-1 and MERS (Middle Eastern Respiratory Syndrome)-CoV. Among the endemic CoV, which have long circulated in the human population, the beta CoV HKU1 and OC43 are more closely related to SARS-CoV-2 than the alpha CoV NL63 and 229E, providing an opportunity to look at the response to a panel of circulating viruses that differ in their degree of similarity. Likewise, we explored binding and functional responses to SARS-CoV-1 and MERS-CoV to examine what breadth may exist for these viruses to which the dyads evaluated here are presumed naïve.

Levels of IgM, IgA, and IgG specific to SARS-CoV-1 but not MERS S protein were elevated in maternal samples from both convalescent and vaccinated dyads as compared to naive subjects (**Figure 5A**). However, different dilutional profiles were observed for these CoV-1 S-specific IgG responses: while infection and vaccination resulted in similar levels of binding antibody detection when serum was tested at a 1:2,500 dilution, when diluted further to 1:5,000, this signal was lost for convalescent subjects while being maintained for vaccinated subjects. As described previously for SARS-CoV-2 antigens, the testing of cord blood samples made clear the detection of low levels of IgM and IgA that react to SARS-CoV-1 and MERS S in even naïve subjects (**Figure 5A**). Similarly, IgM and IgA responses to endemic CoV could be detected in maternal blood by virtue of their absence from cord blood (**Figure 5B**). Whereas vaccination to CoV-2 induced elevated IgG responses to CoV-1 as compared to natural infection when diluted serum samples were tested, the opposite pattern was observed for OC43: infection induced a greater degree of OC43-specific IgG detected at a 1:5,000 dilution, while similar levels were observed when samples were tested at a higher concentration.

**Figure 5.**
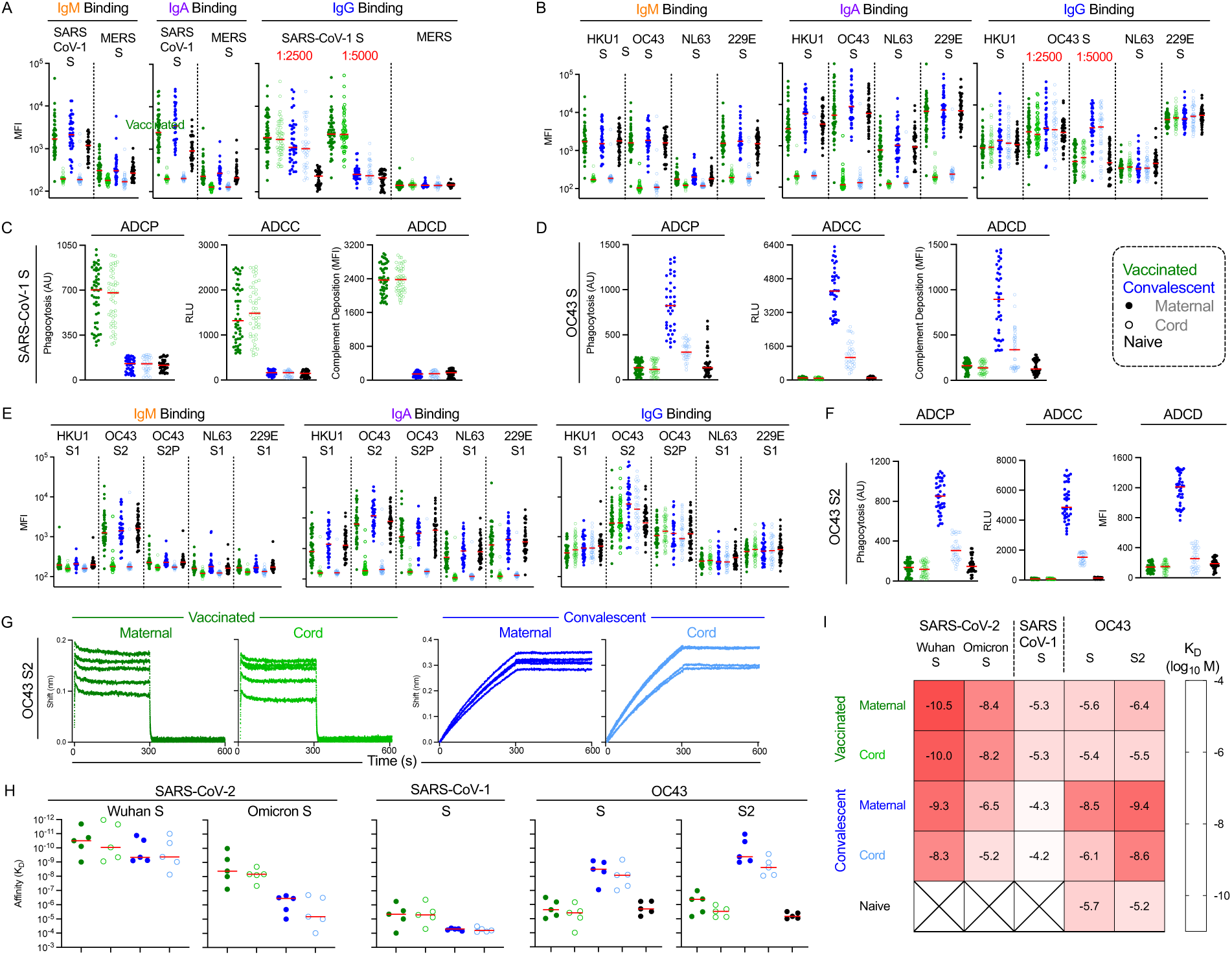
Cross reactivity to emergent and endemic coronavirus. **A.** Coronavirus-specific antibody response features after dimensional reduction in maternal (filled) and cord (open) samples among convalescent (n = 38) (blue) or vaccinated (n = 50) (green) individuals. Naive subjects (n=37) are shown in black. IgG binding experiments were performed at 1:2500 and 1:5000 dilutions. IgM, IgA, and IgG binding responses to SARS-CoV-1 S and MERS S. **B.** IgM, IgA, and IgG binding responses to endemic coronaviruses HKU1, OC43, NL63, and 229E. All antigens are full length spike. **C.** ADCP, ADCC, and ADCD functional responses to SARS-CoV-1 spike. **D.** ADCP, ADCC, and ADCD responses against OC43 S. **E.** IgM, IgA, and IgG antibody binding profiles to endemic coronavirus antigens in other conformations (HKU1 S1, OC43S2, OC43S2P, NL62 S1, 229E S1). **F.** ADCP, ADCC, and ADCD responses against OC43 S2 antigen. Functional activity is reported in arbitrary units (AU), relative light units (RLU), and median fluorescent intensity (MFI). **G.** Antibody association and dissociation traces for affinity analysis of binding to OC43 S2 for a subset of samples from each group (n=5) at 10 mM. **H**. Binding of each subject to the panel of antigens measured. **I.** Heatmap antibody affinities (K_D_ M) to each antigen tested. Darker red denotes higher affinity binding interaction.

Despite the presence of binding antibodies to diverse CoV in both convalescent and vaccinated dyads, antibody effector functions were more limited (**Supplemental Figure 10**). Effector functions were observed in both maternal and cord blood samples of vaccinated but not convalescent dyads for CoV-1 spike (**Figure 5C**), consistent with the binding profiles of dilute serum. These effector functions were present at similar levels in maternal and cord blood, and were lost following IgG depletion (**Supplemental Figure 11**).

Among endemic CoV, vaccinated dyads exhibited only ADCP activity against only stabilized OC43 S2P (**Supplemental Figure 10**), and activity was similar between maternal and cord blood samples. In contrast, convalescent dyads exhibited a diversity of effector functions directed to OC43 S (**Figure 5D**). Consistent with IgG binding across endemic CoV S sub-domains (**Figure 5E**), this activity extended to the OC43 S2 domain (**Figure 5F**), but not to conformationally-stabilized OC43 S2P (**Supplemental Figure 10**). Again, despite the presence of IgM and IgA specific for endemic CoV (**Figure 5B,E**), IgG depletion essentially eliminated all observed activity (**Supplemental Figure 11**).

Due to the distinctions in Fc effector functions and the surprisingly differential dilution-dependence of IgG binding profiles observed for CoV-1 S and OC43 S2 among vaccinated and convalescent samples, we explored if there may be differences in IgG affinity between these groups using biolayer interferometry (BLI) on a subset of samples. Indeed, whereas the total level of binding IgG antibodies was similar to OC43 S2 for vaccinated and convalescent dyads, affinities were distinct among the groups **(Figure 5G-I**). Convalescent dyad samples (n=5) showed binding profiles consistent with higher affinity (multiple orders of magnitude) than the vaccinated dyads, exhibiting the slow on- and slow off-rate profile typical of high affinity interactions. Consistent with functional data, these affinities were lower in convalescent cord than in maternal blood, suggesting a possible explanation for the decreased ADCP, ADCC, and ADCD activity observed in cord blood samples. In contrast, IgG from vaccinated dyads exhibited lower magnitudes and the fast on-, fast off-rate profile commonly seen in low affinity interactions. This latter profile was consistent with that observed in naïve individuals, suggesting that infection induces either recall of pre-existing and/or induction of novel cross-reactive antibodies with high affinity, whereas vaccination does not. Using the same method, we explored whether these affinity differences were specific to OC43 S2 by measuring affinities across a panel of antigens (**Figure 5 H-I**).

Across antigens and serum IgG samples, affinities tended to be marginally lower in cord blood than corresponding maternal samples. Greater differences were observed between vaccinated and convalescent IgG samples. The median affinity of vaccine-elicited IgG binding to Wuhan S exceeded that of IgG induced by natural infection. Relative to Wuhan S, affinity to the Omicron variant was reduced in both dyad groups, however, the reduction was greater for convalescent than vaccinated dyads (**Figure 5 H**). This pattern held with respect to CoV-1 S as well, against which affinity was further decreased. In contrast to emergent CoV, but like the OC43 S2 domain, affinity for OC43 S was greater in convalescent dyads. Vaccinated dyads demonstrated similar affinities for OC43 as were observed in the SARS-CoV-2 naïve subjects. Thus, these affinities corresponded well with effector function profiles: higher effector function and IgG affinity for SARS-CoV-2 antigens were observed in vaccinated dyads, whereas higher effector function and IgG affinity for endemic CoV OC43 antigens were observed in convalescent dyads. These results link effector function to antigen binding affinity as qualitative measures of a polyclonal antibody response that can be distinct from response quantity or magnitude.

## Discussion

The distinct antibody responses to SARS-CoV-2 VOC, endemic, and emergent coronavirus antigens in maternal and cord blood samples from vaccinated and convalescent dyads captured in this study likely result from an array of contributing factors, including antigen conformations, maternal antibody transfer dynamics, affinity differences, and antigen exposure history, among others. A deeper understanding of these factors in the context of pregnancy has the potential to help inform future development of maternal vaccines targeted to contribute to the protection of neonates, particularly as the virus continues to diverge in response to immune pressure mediated by humoral and cellular immunity in the population. Despite the neutralization resistance of VOC, vaccines remain highly effective in preventing severe disease, suggesting the relevance of multiple potential mechanisms including antibody Fc effector functions.

To this end, studying the antibody repertoire of neonates can mimic a passive transfer experiment and has the potential to elucidate some of these mechanisms. Given the persistence and importance of IgG in serum, the selective transfer of maternal IgG makes good biological sense, while the additional passive transfer of maternal IgA present in breastmilk can provide added and more contemporaneous protection at mucosal sites after birth. The evolutionary basis for the preferential transfer of IgGs with certain phenotypes and activities is less clear. Whereas levels of the cytolytic IgG1 subclass in cord blood typically exceed maternal blood, IgG3, arguably the most functionally active subclass, is less well transferred. Transfer of IgG2, relatively inert in terms of effector function is typically also lower, while IgG4 levels appear to be either low enough or sufficiently variable in transfer efficiency that a global trend is less clear^65^. Additionally, while the importance of FcRn in transfer is clear, these patterns don’t precisely recapitulate the serum half-life or *in vitro* binding affinities of the IgG subclasses^66–68^. Further, differences in the efficiency of transfer of antibodies with different pathogen- or antigen-specificities^24,69^ , as well as with variable glycosylation^70–72^, particularly in the variable region^70^, are also associated with differences in transfer efficiency. However, the means by which these attributes may contribute rather than simply correlate with other, mechanistically relevant factors is unclear. The longitudinal profile of maternal responses and total serum IgG levels clearly each play a role^73–75^, but samples are typically only tested at a snapshot in time, often at delivery, as opposed to characterized with more continuous kinetic profiling. Recent work has raised the possibility that additional processes or transporters, including FcγR, may play a role^44,46^.

Here, by comparing the SARS-CoV-2 spike-specific antibodies with those that recognize non-sarbecovirus antigens, we assessed some of these factors. The generally poorer transfer of total and each individual IgG subclass for SARS-reactive antibodies as compared to antigens that mothers were presumed to be seropositive against before pregnancy suggests the importance of their more recent induction and the contribution of the length of time over which maternal antibodies can be transferred in the levels observed in neonates. In contrast, the general consistency between well and poorly transferred subclasses, independent of specificity, points to this factor as generalizable. The relative efficiency of SARS-CoV-2-specific antibody transfer between dyads with a history of vaccination as compared to infection also differed somewhat, with generally improved transfer observed in vaccinated dyads, consistent with a possible influence of inflammation associated with maternal infection.

Antibody functions were also variably well transferred between exposure history groups. ADCP, ADCC, and ADCD activities against pertussis and tetanus antigens were generally elevated in cord blood, while Wuhan SARS-CoV-2 spike-specific antibody effector functions were typically similar in vaccinated and slightly reduced in convalescent cord as compared to maternal blood. For variants, however, this decrease was dramatic in the context of natural infection, and some activities were essentially undetectable in convalescent cord blood. The reduction in the breadth of antibody function paralleled the reduced breadth of antibody binding and activity observed in natural infection overall, but the magnitude of functional loss suggests that non-linearity, or threshold effects are at play in these activities. As has been reported elsewhere, this pattern was also apparent in neutralization activity, which was greatly reduced in cord blood. As compared to effector functions, broad neutralization activity was relatively sparse, even in maternal blood. Among convalescent participants, even dyads with high Wuhan strain neutralizing titers (>1:200) in maternal serum typically exhibited undetectable activity in cord blood. The poorer neutralization activity of antibodies in cord blood than maternal serum is likely at least partly attributable to the loss of contributions from IgM and IgA. Among other possible contributing factors, this study identified differences in the affinity for antigen as potentially playing a role in defining breadth across effector functions. Vaccination led to higher affinity antibodies against emergent coronaviruses whereas natural infection induced higher affinity antibodies towards endemic coronaviruses. In turn, these affinity profiles were consistent with antibody effector function breadth and potency profiles. Relative to IgG1, the poorer transfer of IgG3, which can exhibit both greater effector function and greater apparent affinity for antigen associated with its greater flexibility and hinge length, may also account for some of the differences observed between maternal and cord blood and between vaccination and infection.

Consistent with prior reports^76^, differences were observed in the relative levels of responses and effector functions of SARS-CoV-2- and the endemic CoV OC43-specific antibodies that are associated with spike stabilization and differences in cross-reactivity of antibodies directed to different sub-domains. However, this study associates these functional distinctions with antibody affinity for antigen, finding that polyclonal IgG pools with higher affinity antibodies exhibit greater effector function. While this study cannot address the relative importance of one antibody function over another, neutralization activity was considerably more limited than binding and effector functions, with less than one in ten dyads exhibiting detectable neutralization of the Omicron strain. Low levels of binding antibodies to other emergent and endemic CoV could be detected, and some were present at levels and with characteristics, such as high affinity, sufficient to induce effector functions. Domain-level mapping of responses directed to endemic CoV implicated the highly conserved S2 domain in these exceptionally broad responses, particularly in association with natural infection, and prior work has suggested that they result from cross-reactive clones^76,77^. As VOC continue to emerge, and antigen-experienced populations benefit from mucosal antibody and T cell responses, the relative importance of serum antibody neutralization titers, which have served as a robust correlate of protection in early efficacy trials^78–82^, may vary. Indeed, the mechanistic relevance of neutralization, at least as typically tested *in vitro*, has been challenged by observations that non-neutralizing and even antibodies that increase viral infectivity *in vitro* can provide protection *in vivo* ^83,84^. To this end, the broad recognition and function of antibodies raised by vaccination and natural infection support the feasibility of “universal” COVID-19 vaccine development efforts.

Some limitations this study have already been alluded to. Maternal antibodies were sampled only at the time of delivery, so their dynamic profile is not known. Further, the impact of timing of maternal seropositivity could not be meaningfully evaluated given the relatively narrow window during gestational ages at which exposure occurred. While study participants were enrolled at a similar timepoints in the pandemic, they were drawn from geographically distinct populations. With the exception of neutralization, antibody functions were evaluated in simplified rather than more ideally biologically authentic assays, and cell lines rather than neonatal effector cells were employed to characterize activity in cord blood.

Sufficient sample volumes were not available to support evaluation of all specificities and activities in all assays. The mechanisms whereby binding and functional assays show different transfer efficiencies are likely to relate at least in part to differences in IgG subclass transfer, but further study would be needed to more clearly resolve the roles of each antibody attribute to the efficiency of transfer and to each function. Further, evidence of threshold effects was observed in several assays, but it remains unclear whether the thresholds observed in the *in vitro* assays employed here are consistent those that might exist *in vivo*.

Lastly, while the relevance of antibody binding and effector functions tested in depth here to protection from disease has been suggested in many prior studies^85^, insights into the infection resilience of either mothers or neonates in this study is lacking, and therefore relationships between these measures and disease cannot be addressed here.

Overall, while numerous studies have shown the reduction in neutralization following viral diversification over time, vaccines remain highly effective at preventing severe disease and death, pointing to the contributions of other immune mechanisms. This study addresses the passive transfer and inter-generational inheritance of functionally potent antibodies, and the relative ability of IgG antibodies to drive broad recognition and effector activity that may contribute to protection from COVID-19. While SARS-CoV-2-specific neutralizing activity was typically lost and antibody binding breadth and effector function were typically reduced in cord as compared to maternal blood, effector functions were substantially greater and broader following maternal vaccination than infection in both mothers and infants. The biases in levels, isotypes, subclasses, affinity for antigen, neutralization and effector function breadth and potency that associate with antigen exposure history have implications for protecting diverse populations from ever-diversifying viral variants.

## Methods

### Human Subjects

Vaccinated participants (Israel), screened for lack of anti-N SARS-CoV-2 antibody responses, received two doses of mRNA-encoded stabilized spike BNT162b2 (n=50) vaccine. Convalescent participants (Belgium), had infection status defined by RT-PCR (n=38). While Wuhan was the dominant strain in circulation at the time of sample collection, viruses were not typed. Naïve serum was obtained from a commercial vendor (BioIVT) prior to approval of vaccines and was screened for anti-N SARS-CoV-2 antibody responses to exclude donors with previous infection. Characteristics for each study group are described in **Supplemental Table 1**. While pregnant subjects completed their vaccination series in the third trimester, and most convalescent subjects reported symptoms or tested positive in their third trimester, elapsed time since most recent SARS-CoV-2 antigen exposure differed between cohorts, as did time to delivery following diagnosis or receipt of the second vaccine dose. Study participants provided informed written consent and studies were reviewed and approved by IRBs at individual collection sites and Dartmouth.

### Fc Array

Antigens were purchased from commercial sources or transiently expressed in Expi293 or HEK293 cells and purified via affinity chromatography (**Supplemental Table 2**). Fc receptors were expressed and purified as described previously^86^. Antigen-specific antibodies were characterized using the Fc array assay^85,87^. Briefly, antigens were covalently coupled to MagPlex microspheres (Luminex Corporation).

Experimental controls included pooled human polyclonal serum IgG (IVIG), S309 an antibody from a SARS-CoV patient that cross-reacts SARS-CoV and SARS-CoV-2, and VRC01, an HIV specific antibody^88,89^. Serum dilutions for profiling varied from 1:250 to 1:5000 depending on detection reagent. Unless otherwise noted, concentrations tested were as listed in **Supplemental Table 2**. Antigen-specific antibodies were detected by R-phycoerythrin-conjugated secondary reagents specific to human immunoglobulin isotypes and subclasses and by Fc receptor tetramers^90,91^. Median fluorescent intensity data was acquired on a FlexMap 3D array reader (Luminex Corporation). Samples were run in technical duplicate.

### Neutralization

SARS-CoV-2 neutralizing antibodies (nAb) were quantified as previously reported^85,92^ for a subset of maternal and matched cord blood serum samples (23 vaccinated, 26 convalescent, selected based on having the highest binding antibody levels from among dyads with sufficient serum volumes available). Briefly, serial dilutions of heat-inactivated serum (1/50 to 1/25,600 in EMEM supplemented with 2 mM L-glutamine, 100 U/ml-100ug/mL of Pencillin-Streptomycin and 2% fetal bovine serum) were incubated for 1 hr at 37°C and 7% CO_2_ with 3xTCID_100_ of Wuhan strain (2019-nCoV-Italy-INMI1, 008 V-03893) and Omicron strain BA.1 (B1.1.529, VLD20211207). A volume of 100 μL of sample-virus mixture was added to 100 μL of Vero cells (18,000 cells/well) in a 96 well plate and cultured for five days at 37°C and 7% CO_2_. Cytopathic effects of viral growth were scored microscopically and the Reed-Muench method was used to calculate the nAb titer that reduced the number of infected cells by 50% (NT_50_), which was used as a proxy for the nAb concentration in the sample. An internal reference standard composed of a pool of serum from naturally infected and vaccinated adults was included in each nAb assay run, which was calibrated against the Internationals Standard 21/234 (NIBSC), in accordance with WHO guidance.

### Antibody-Dependent Cellular Phagocytosis (ADCP)

Characterization of the phagocytic activity of serum antibodies was performed as described previously^85,93^. Briefly, 1 μM yellow-green fluorescent beads (Thermo Fisher, F8813) were covalently conjugated to antigen. Beads were then incubated with serum samples for 4 hr with THP-1 cells (ATCC TIB-202) at 37°C in 5% CO_2_. Afterwards, cells were fixed and analyzed by flow cytometry using a MACSQuant Analyzer (Miltenyi Biotec) to define the percentage of cells that phagocytosed one or more fluorescent beads and the MFI of this population, the product of which was defined as the phagocytic score (arbitrary units). Controls included wells with no added antibody were used to determine the level of antibody-independent phagocytosis, S309 and VRC01 antibodies, and concentrated pooled polyclonal serum IgG (Sigma Aldrich I4506); samples were tested in three biological replicates.

### Surrogate Reporter Cell Assay of Antibody-Dependent Cellular Cytotoxicity (ADCC)

A CD16 activation reporter assay system was used as a surrogate for ADCC^85,94^. First, high binding 96-well plates were coated overnight at 4⁰C with 1 μg/mL of spike or RBD antigen. Following incubation, plates were washed (PBS + 0.1% Tween20) and blocked (PBS + 2.5% BSA) at room temperature (RT) for 1 hr. Following plate washing, 100,000 CD16- (FcγRIIIa) expressing Jurkat Lucia NFAT (Invivogen, jktl-nfat-cd16) cells, cultured according to manufacturer’s instructions, and dilute serum samples were added to each well in cell culture media lacking antibiotics in a 200 μL volume. After 24 hrs of culture, 25 μL of supernatant from each well was transferred into a white 96 well plate to which 75 μL of quantiluc substrate was added. After 10 min, luciferase signal was determined by reading plates on a SpectraMax plate reader (Molecular Devices). Assay controls included cell stimulation cocktail (Thermo Fischer Scientific, 00-4970-93) and ionomycin, buffers alone, spike-specific S309 and HIV-specific VRC01 monoclonal antibodies, and concentrated pooled polyclonal serum IgG (Sigma Aldrich I4506). Samples were run in three biological replicates. In a prior study, this assay correlated well with killing activity against a spike-expressing cell line^85^.

### Antibody-Dependent Complement Deposition (ADCD)

Antibody-dependent complement deposition (ADCD) experiments were performed essentially as previously described^85,95^. Serum samples were heat inactivated for 30 min at 56°C, prior to incubation for 2 hr at RT with assay microspheres. Human complement serum (Sigma, S1764) diluted 1:100 in gel veronal buffer (Sigma-Aldrich, GVB++, G6514) was mixed with samples and microspheres at RT with shaking for 1 hr. After washing, samples were incubated with murine anti-C3b (Cedarlane #CL7636AP) at RT for 1 hr followed by staining with anti-mouse IgG1-PE secondary Ab (Southern Biotech #1070-09) at RT for 30 min. A final wash was performed and samples were resuspended into Luminex sheath fluid and MFI acquired on a FlexMap 3D reader. Assay controls included heat-inactivated complement, buffers alone, spike-specific S309 and HIV-specific VRC01 monoclonal antibodies, and concentrated pooled polyclonal serum IgG (Sigma Aldrich I4506). Samples were run in three biological replicates.

### Antibody depletion

Antibody depletion experiments were performed on a subset of samples (n=15). Depletions were performed using Ig capture select resins following manufacturer’s instructions (Thermo Fisher) to deplete IgM, IgA, or IgG. Depleted samples along with a mock control were tested in the multiplex assay to measure depletion efficiency. Mock depletion samples followed the same protocol as the manufacturer’s instructions with the exception being that an irrelevant column matrix (Ni-NTA Thermo Fisher) was used.

### Octet Analysis

Serum was purified for IgG using Melon Gel Purification kit (Thermo Fisher) following the manufacturer’s instructions. Binding affinities were determined using biolayer interferometry (BLI) on the Forte Bio Octet system, essentially as previously described^96^. Antigens were biotinylated with LC-LC no weigh biotin (Thermo Fisher, A39257). After 30 min of reaction time, excess biotin was removed with Zeba desalting columns (Thermo Fisher, 89882). To determine binding kinetics, biotinylated antigens were captured on streptavidin Sax-2.0 tips (Forte Bio, 18-5136) and then incubated in dilute IgG. Briefly, biosensors were first equilibrated in PBST (0.05% Tween-PBS) for 180s and activated by dipping into 10 mM glycine (pH 1.7) for 20s and PBST 20s for three cycles. Biosensors were then loaded with biotinylated antigen at 1mg/ml in PBST for 300s, and dipped into PBST for 300s to reach baseline, prior to a 300s association phase in which they were dipped into IgG, and a 300s dissociation phase in which they were dipped into PBST. Assessments were performed across 3-fold serial dilutions of IgG ranging from 10 mM to 0.013 mM. Sample traces of a single concentration (10mM) were plotted for representative results. Tips were regenerated for 20s for each condition in 10mM glycine pH 1.7. Data was aligned and corrected between steps as needed, and signal observed in reference sample wells, comprised of tips loaded with antigen but not dipped into IgG, was subtracted. For kinetic analysis, a 1:1 association and dissociation model was selected in Forte Bio data analysis 7.0 software in order to determine K_D_.

### Data Analysis and Statistical Quantification

UMAP plots were generated in Python (version 3.11) using the umap-learn package (version0.4). Volcano plots were generated in R (version 4.3) using ggplot2. Statistical analysis was performed in GraphPad Prism (version 9.7). Statistical test are described in the respective figure legends. Breadth-potency curves were defined as the proportion of antigen-specificities exhibiting a signal above a given intensity. Curves were generated using the LOWESS curve fit method in Prism for each respective subject group. Breadth scores were calculated by taking the geometric mean across antigen specificities for each subject. The sample size for each figure includes all subjects from their respective groups unless otherwise noted.

## Data Availability

All data produced in the present study are available upon reasonable request to the authors.

## Acknowledgements

We would like to thank all participants who enrolled in this study and the study and laboratory staff who helped collect and process the samples. A number of antigen expression constructs were provided by Dr. Jason McLellan (UT Austin) and the positive control mAb S309 was provided by Dr. Jiwon Lee (Dartmouth). The following reagent was produced under HHSN272201400008C and obtained through BEI Resources, NIAID, NIH: Spike Glycoprotein Receptor Binding Domain (RBD) from SARS-Related Coronavirus 2, Wuhan-Hu-1 with C-Terminal Histidine Tag, Recombinant from Baculovirus, NR-52307. The following reagent was deposited by the Centers for Disease Control and Prevention and obtained through BEI Resources, NIAID, NIH: SARS-Related Coronavirus 2, Isolate USA-WA1/2020, NR-52281.

## Conflict of Interest

M.E.A. reports grant support from Be Bio and Moderna unrelated to COVID-19 vaccines.

## Funding

This work was supported in part by the National Institute of Allergy and Infectious Diseases 1U19AI145825, National Cancer Institute P30 CA 023108-41, National Institute of General Medical Sciences P20-GM113132. A.M. is Research Director at the F.R.S., FNRS, Belgium.

## Notes

### Author Declarations

IRBs at CHU St. Pierre, Hadassah Medical Center,BioIVT clinical sites and Dartmouth gave ethical approval for this work.

