## Supplemental Materials for "Patterns and functional consequences of antibody speciation in maternal-fetal transfer of coronavirus-specific humoral immunity"

**Supplementary Materials**

| **Supplementary Figures** | |
|---|---|
| Supplementary Figure 1 | IgM, IgA, and IgG responses to SARS-CoV-2 VOC RBD antigens |
| Supplementary Figure 2 | IgG subclass binding to spike and RBD antigens |
| Supplementary Figure 3 | FcγR binding to SARS-CoV-2 variant spike and RBD-specific antibodies |
| Supplementary Figure 4 | IgG subclass and FcγR-binding antibody transfer ratios for SARS-CoV-2 RBD antigens |
| Supplementary Figure 5 | Functional responses for cord and maternal samples to SARS-CoV-2 spike antigens. |
| Supplementary Figure 6 | Functional responses for cord and maternal samples to SARS-CoV-2 RBD antigens. |
| Supplementary Figure 7 | Fc effector function transfer ratios for activity toward SARS-CoV-2 RBD antigens |
| Supplementary Figure 8 | Changes in Ig isotype levels following depletion |
| Supplementary Figure 9 | Ig isotype specificity of depletion procedure for each tested sample |
| Supplementary Figure 10 | Functional responses to emergent and endemic coronaviruses |
| Supplementary Figure 11 | Effect of IgG depletion on effector function observed for SARS-CoV-1 and OC43 S. |
| **Supplementary Tables** | |
| Supplementary Table 1 | Subject Cohorts |
| Supplementary Table 2 | Fc Array reagents and test conditions |

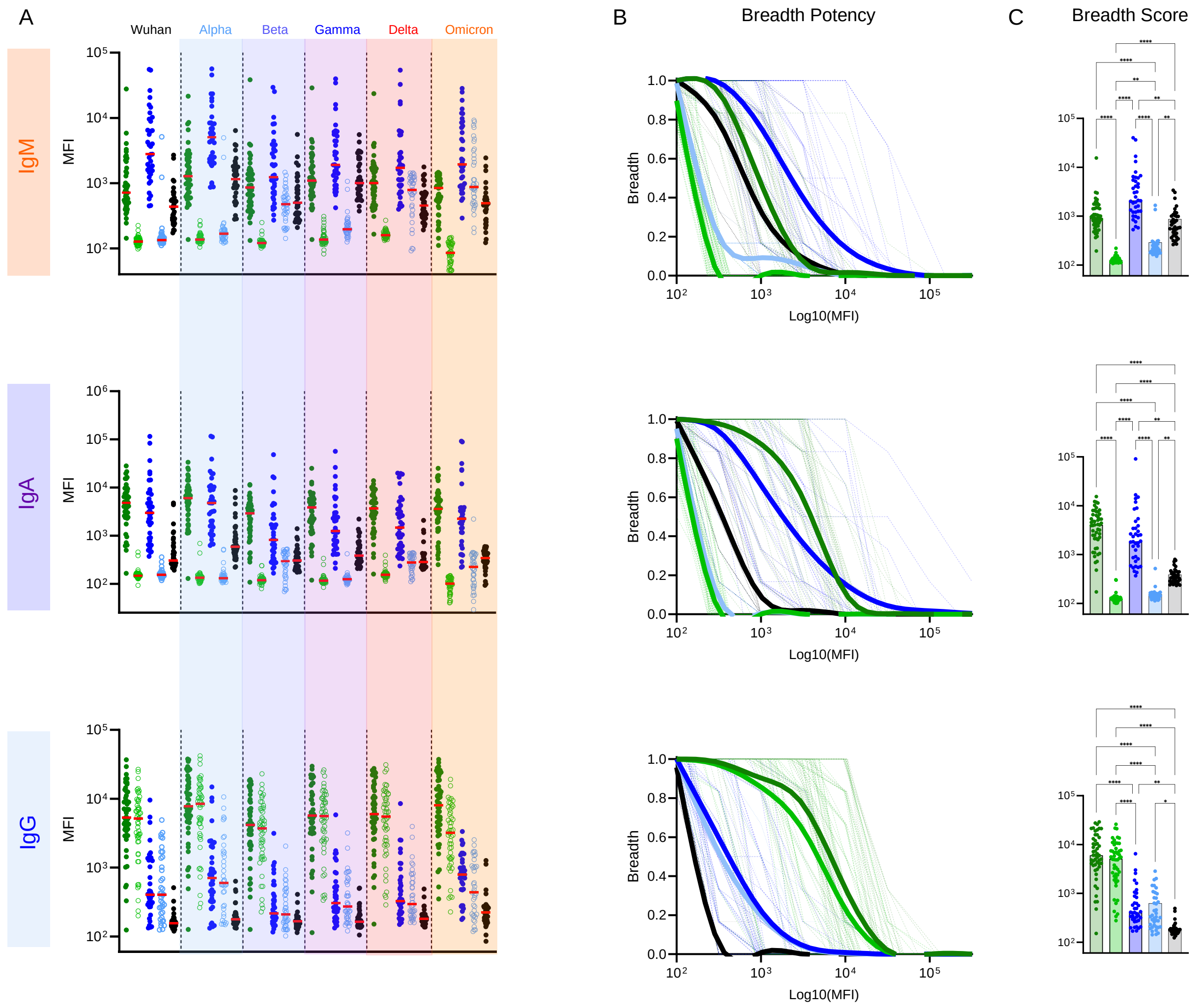

**Supplementary Figure 1. IgM, IgA, and IgG antibody responses to SARS-CoV-2 VOC RBD antigens. A.** IgM, IgA, and IgM antibody binding responses in maternal (filled) and cord (open) samples among convalescent (n = 38) (blue) or vaccinated (n = 50) (green) individuals against SARS-CoV-2 RBD antigens. Naïve subjects (n=37) are shown in black. **B.** Breadth-potency curves represent the fraction of subjects with a response exceeding a given level for IgM, IgA, and IgG antibody responses across the panel of VOC. Population means are shown with a thick line, and individual subjects illustrated in thin lines. **C.** IgM, IgA, and IgG breadth scores for each subject. Bar indicates the median. Statistical significance was defined by ANOVA Kruskal–Wallis test with Dunn's correction and α=0.05 (*p<0.05, **p<0.01, ***p<0.001, ****p<0.0001).

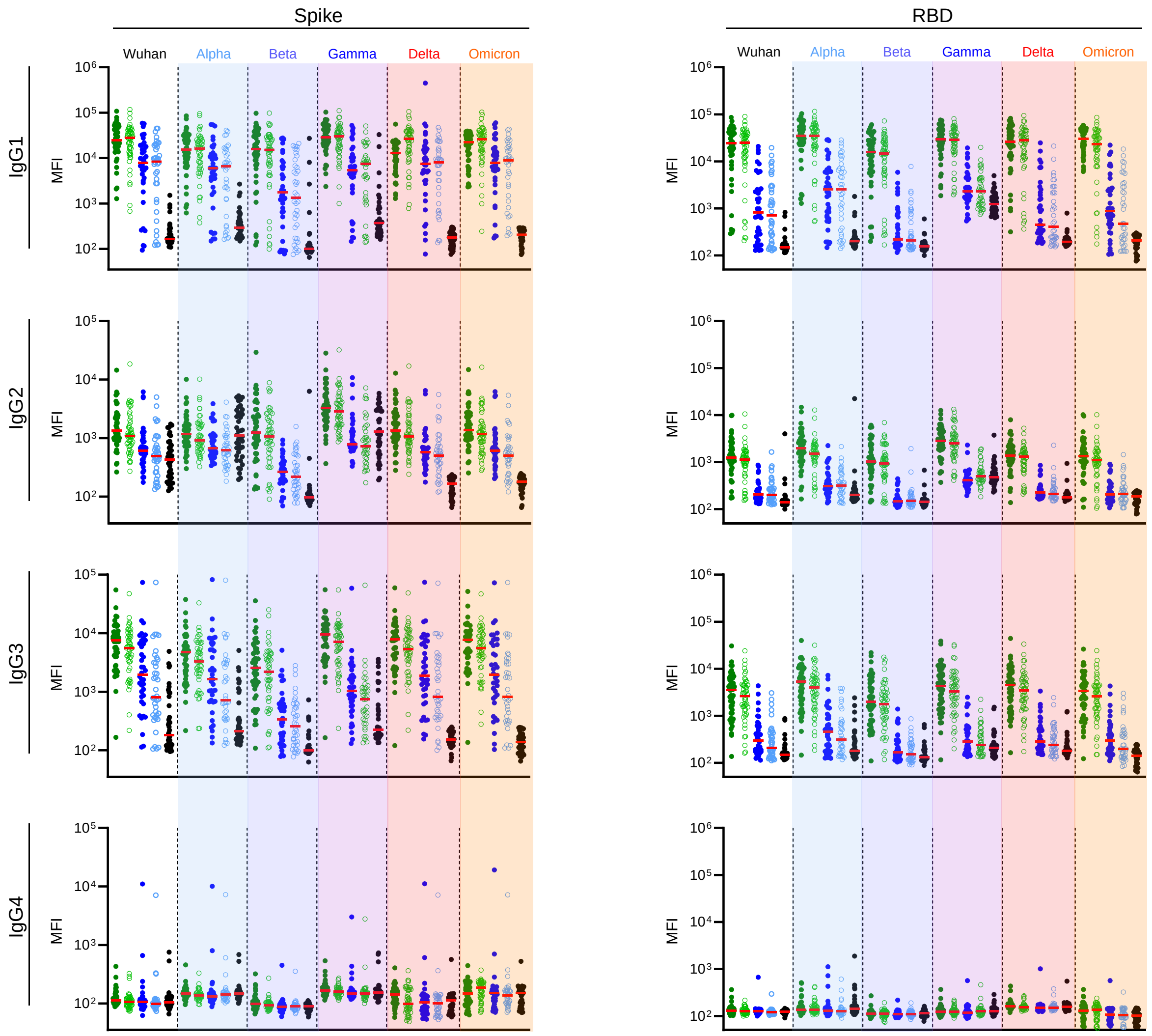

**Supplementary Figure 2. IgG subclass binding to SARS-CoV-2 variant spike and RBD antigens.** IgG subclasses binding activities toward spike and RBD-specific antibodies across Wuhan and VOC proteins in serum for maternal (filled) and cord (open) samples among convalescent (n = 38) (blue) or vaccinated (n = 50) (green) individuals. Naïve (n=37) (black) subjects. Bar indicates median.

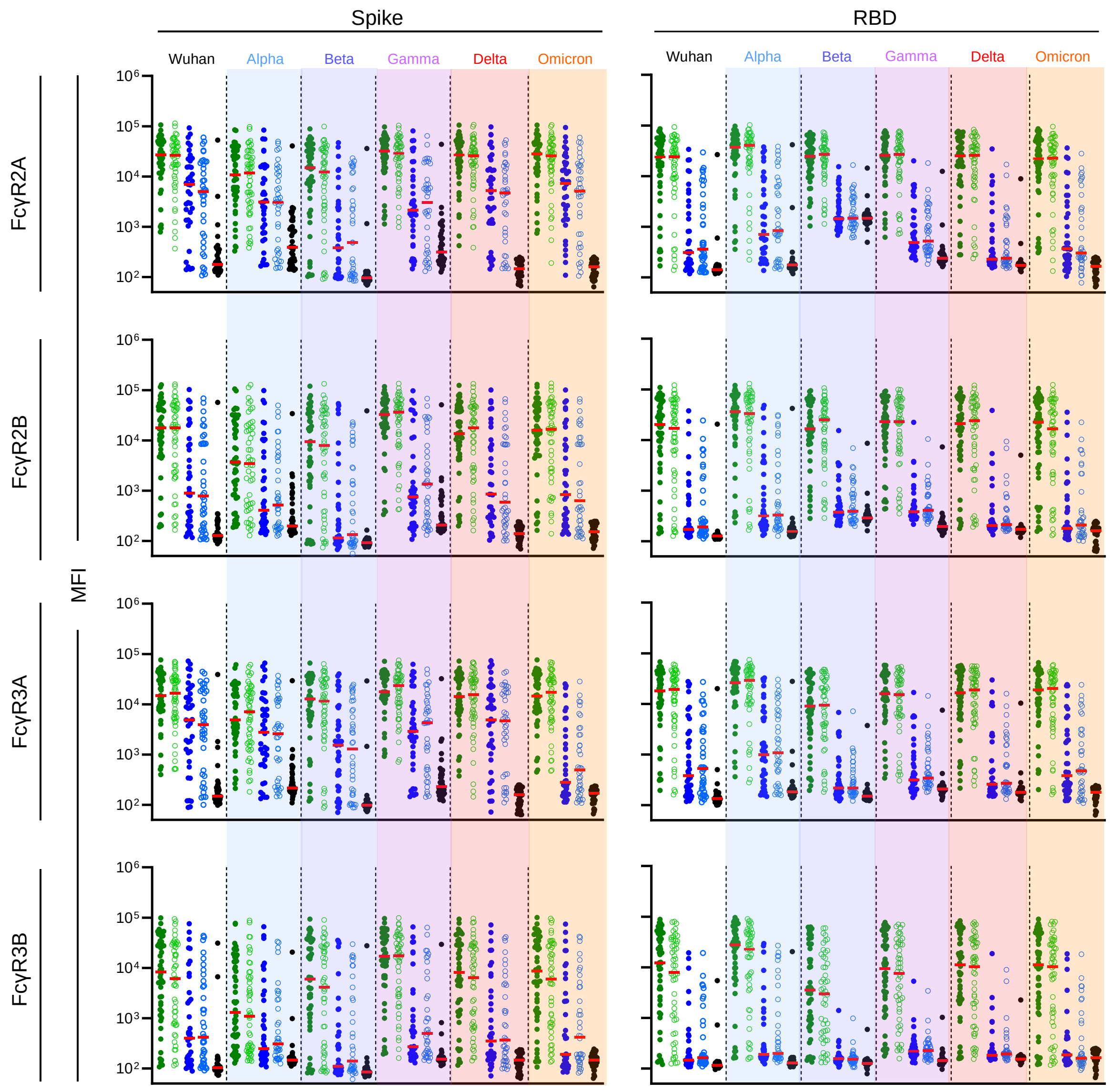

**Supplementary Figure 3. FcγR binding to SARS-CoV-2 variant spike and RBD-specific antibodies.** Fc receptor binding activities of spike- (left) and RBD- (right) specific antibodies across Wuhan and VOC proteins in serum in maternal (filled) and cord (open) samples among convalescent (n = 38) (blue) or vaccinated (n = 50) (green) individuals. Naïve (n=37) (black) subjects. Bar indicates median.

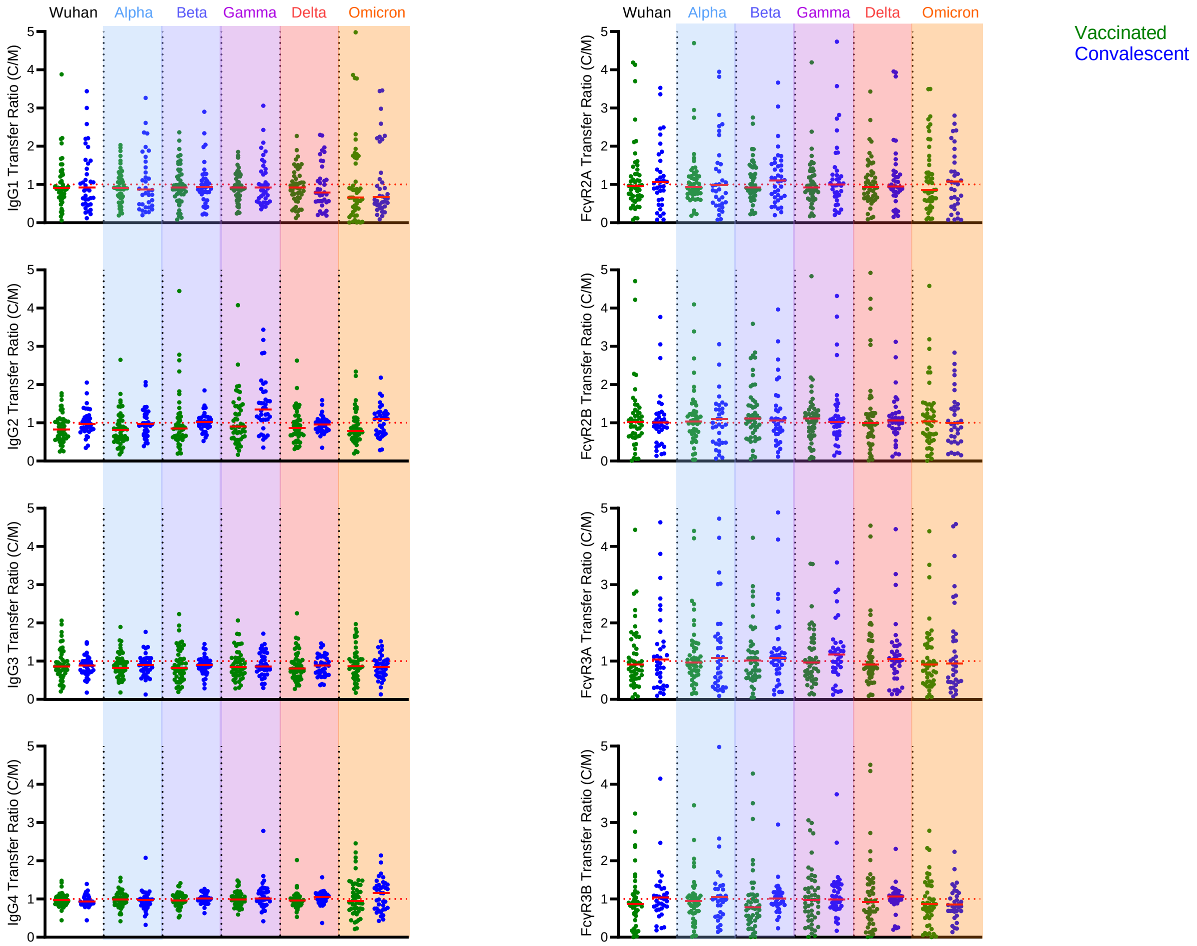

**Supplemental Figure 4. IgG subclass and FcγR-binding antibody transfer ratios for SARS-CoV-2 RBD antigens.** Transfer ratio (cord/maternal levels) SARS-CoV-2 RBD-specific IgG subclasses (left) and Fc receptor antibody binding (right) in vaccinated (green) and convalescent (blue) dyads. Bar indicates median.

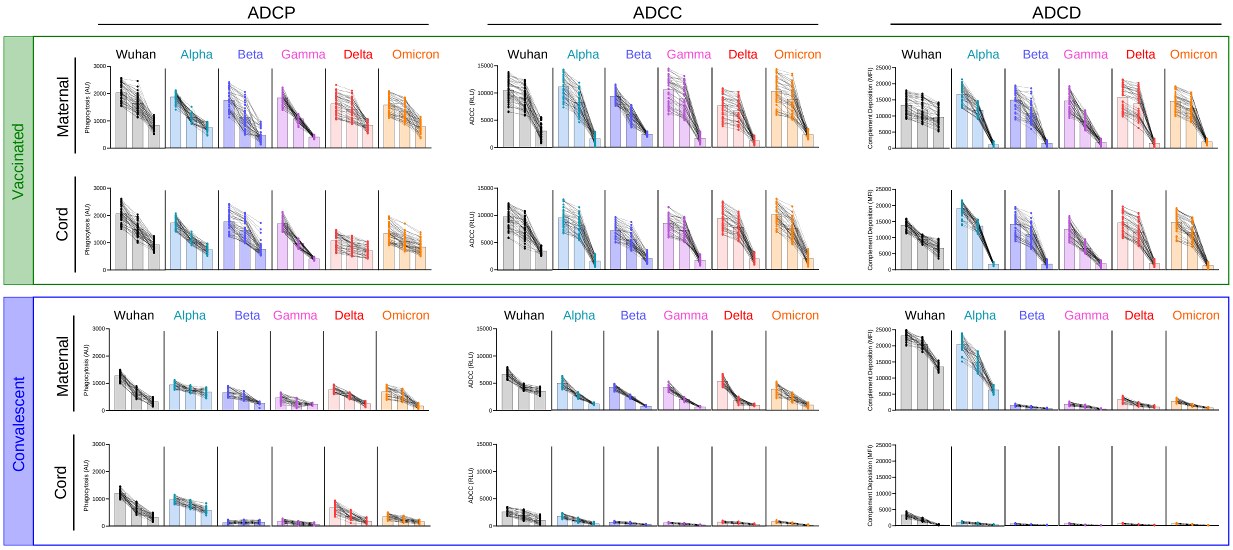

**Supplementary Figure 5. Functional responses for cord and maternal samples to SARS-CoV-2 spike antigens.** Ab effector functions from vaccinated maternal and cord blood (top, green box) and convalescent maternal and cord blood (bottom, blue box) for SARS-CoV-2 spike variants. Phagocytosis (left), ADCC (center), and Complement Deposition (ADCD, right) activities were assessed at each of three serum dilutions (1:50, 1:100, 1:250). Individual traces for each subject across dilutions are displayed. Functional activity is reported in arbitrary units (AU), relative light units (RLU), and median fluorescent intensity (MFI). Bar indicates median.

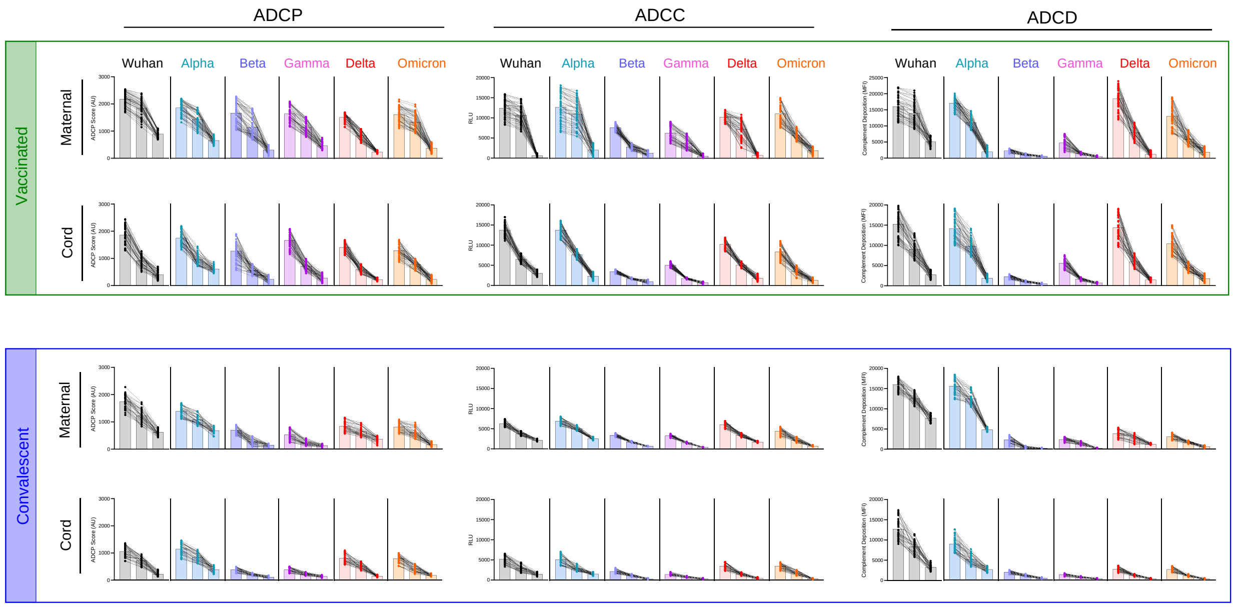

**Supplementary Figure 6. Functional responses for cord and maternal samples to SARS-CoV-2 RBD antigens.** Ab effector functions from vaccinated maternal and cord blood (top, green box) and convalescent maternal and cord blood (bottom, blue box) for SARS-CoV-2 RBD variants. Phagocytosis (ADCP, left), ADCC (center), and Complement deposition (ADCD, left) activities were assessed at each of three serum dilutions (1:50, 1:100, 1:250). Individual traces for each subject across dilutions are displayed. Functional activity is reported in arbitrary units (AU), relative light units (RLU), and median fluorescent intensity (MFI), respectively. Bar indicates median.

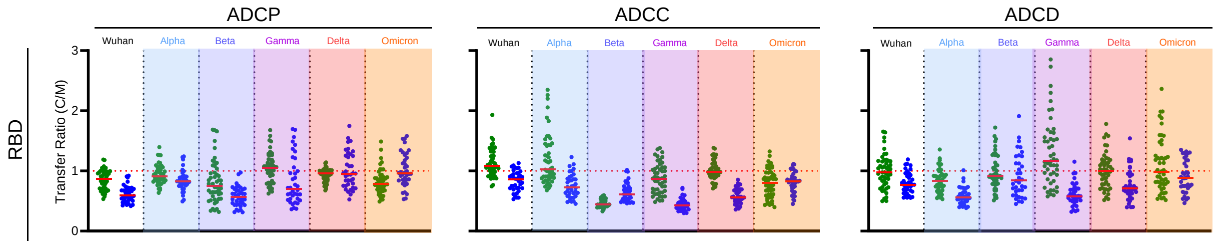

**Supplementary Figure 7. Fc effector function transfer ratios for activity toward SARS-CoV-2 RBD antigens.** Transfer ratio (cord/maternal levels) of antigen-specific (SARS-CoV-2 VOC RBD antigens) Fc effector functions including ADCP (left), ADCC (center) and ADCD (right) in vaccinated (green) and convalescent (blue) dyads. Bar indicates median.

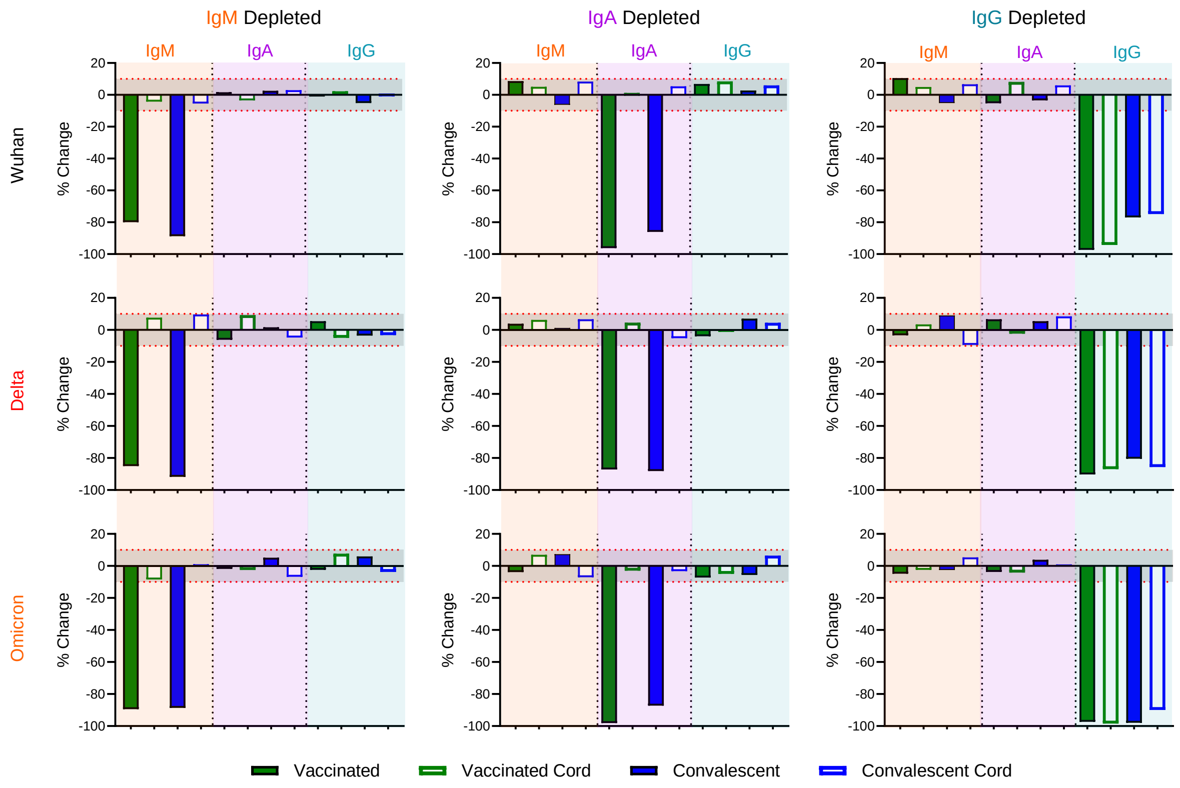

**Supplemental Figure 8. Changes in Ig isotype levels following depletion.** Maternal (filled) and cord (open) samples among convalescent (n = 15) (blue) or vaccinated (n = 15) (green) individuals were depleted of IgM (left), IgA (center), or IgG (right). Detection of antigen-specific immunoglobulin of each isotype was measured for each sample and percent change was calculated based on a mock control for each respective sample. Depletions were measured for antigen-specific antibodies to Wuhan (top), Delta (center), and Omicron (bottom) spike antigens.

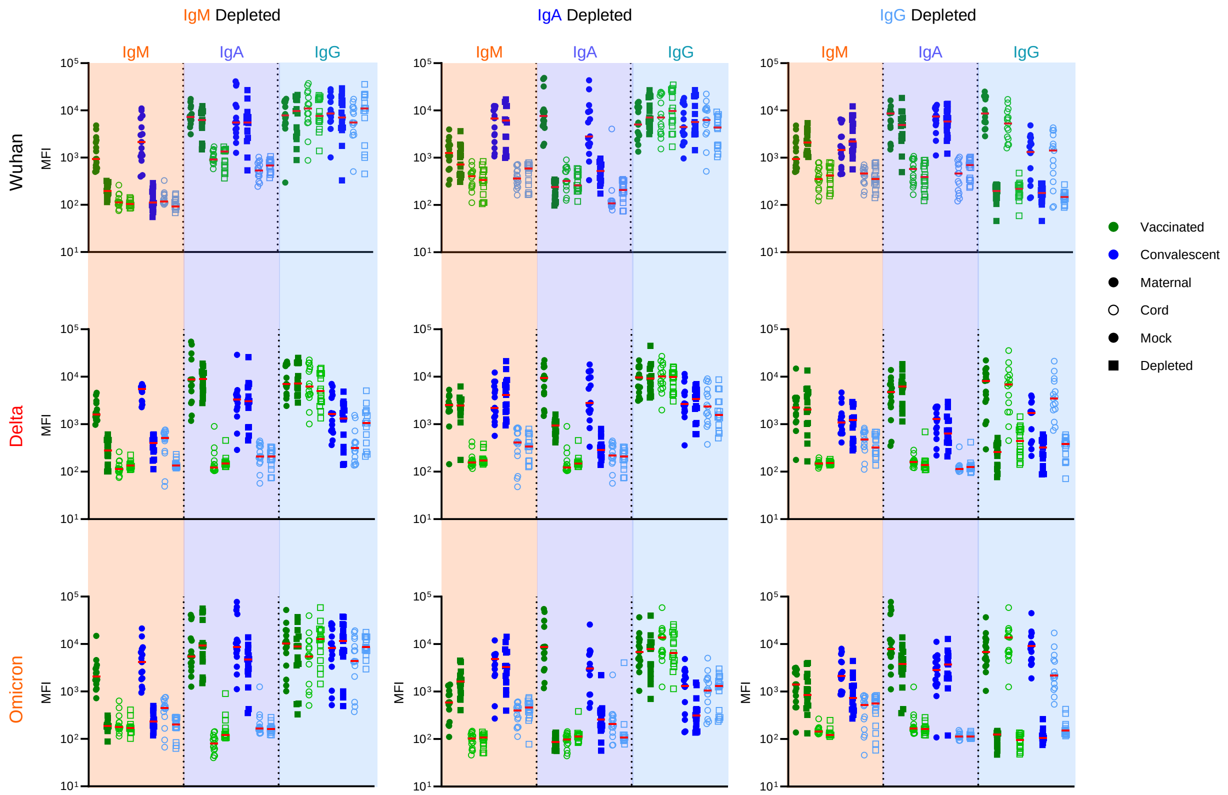

**Supplemental Figure 9. Ig isotype specificity of depletion procedure for each tested sample.** Maternal (filled) and cord (open) samples among convalescent (n = 15) (blue) or vaccinated (n = 15) (green) individuals were depleted of IgM, IgA, or IgG. Circle and square shapes denote mock and depleted samples, respectively. Binding was measured for each sample. Depletions were measured for antigen specific antibodies to Wuhan (top), Delta (center), and Omicron (bottom) spike antigens. Bar indicates median.

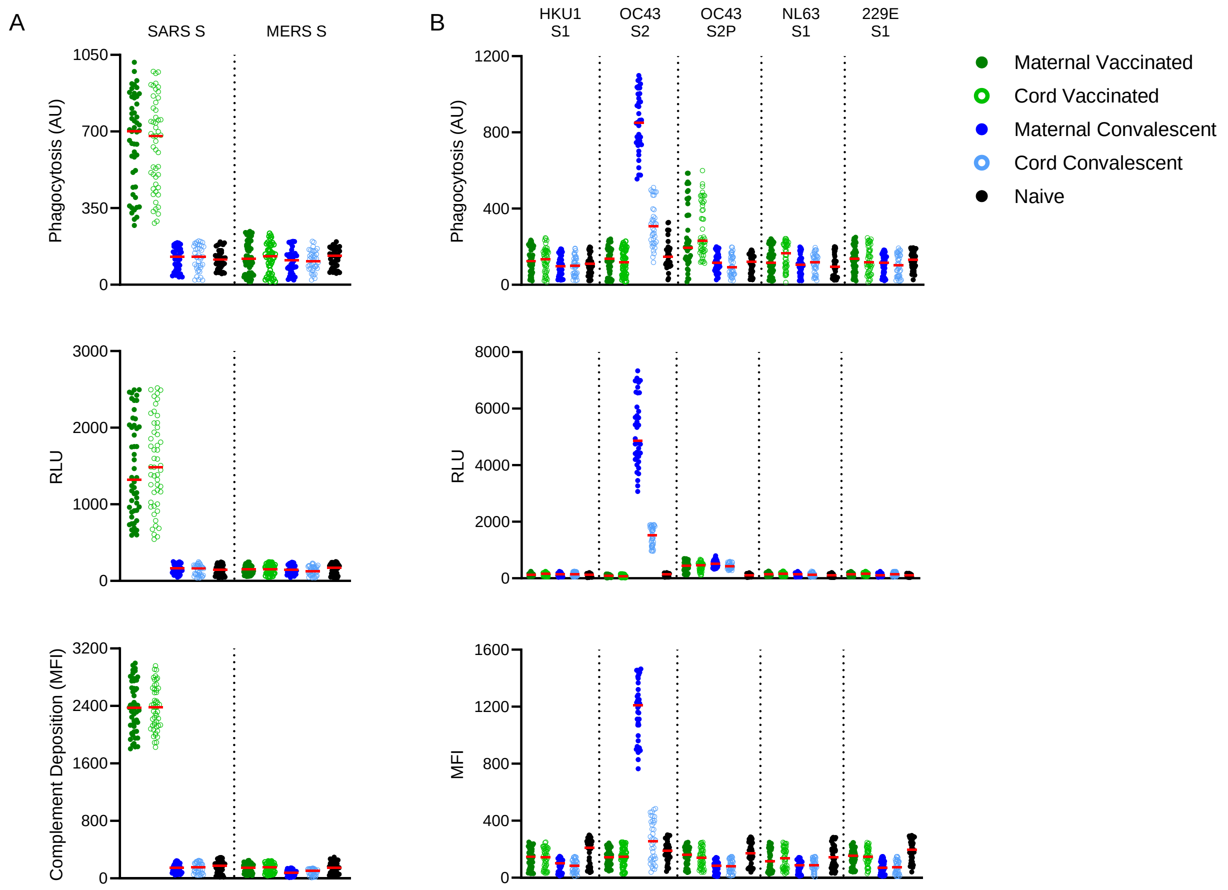

**Supplemental Figure 10. Functional responses to emergent and endemic coronaviruses.** Fc effector functions against emergent coronaviruses SARS-CoV-1 and MERS-CoV (**A**) and endemic coronaviruses (**B**) HKU1, 0C43, NL63, and 229E. Maternal (filled) and cord (open) samples among convalescent (n = 38) (blue) or vaccinated (n = 50) (green) individuals were tested for ADCP (top), ADCC (center), and ADCD (bottom). Naïve subjects are shown in black (n=37). Bar indicates median.

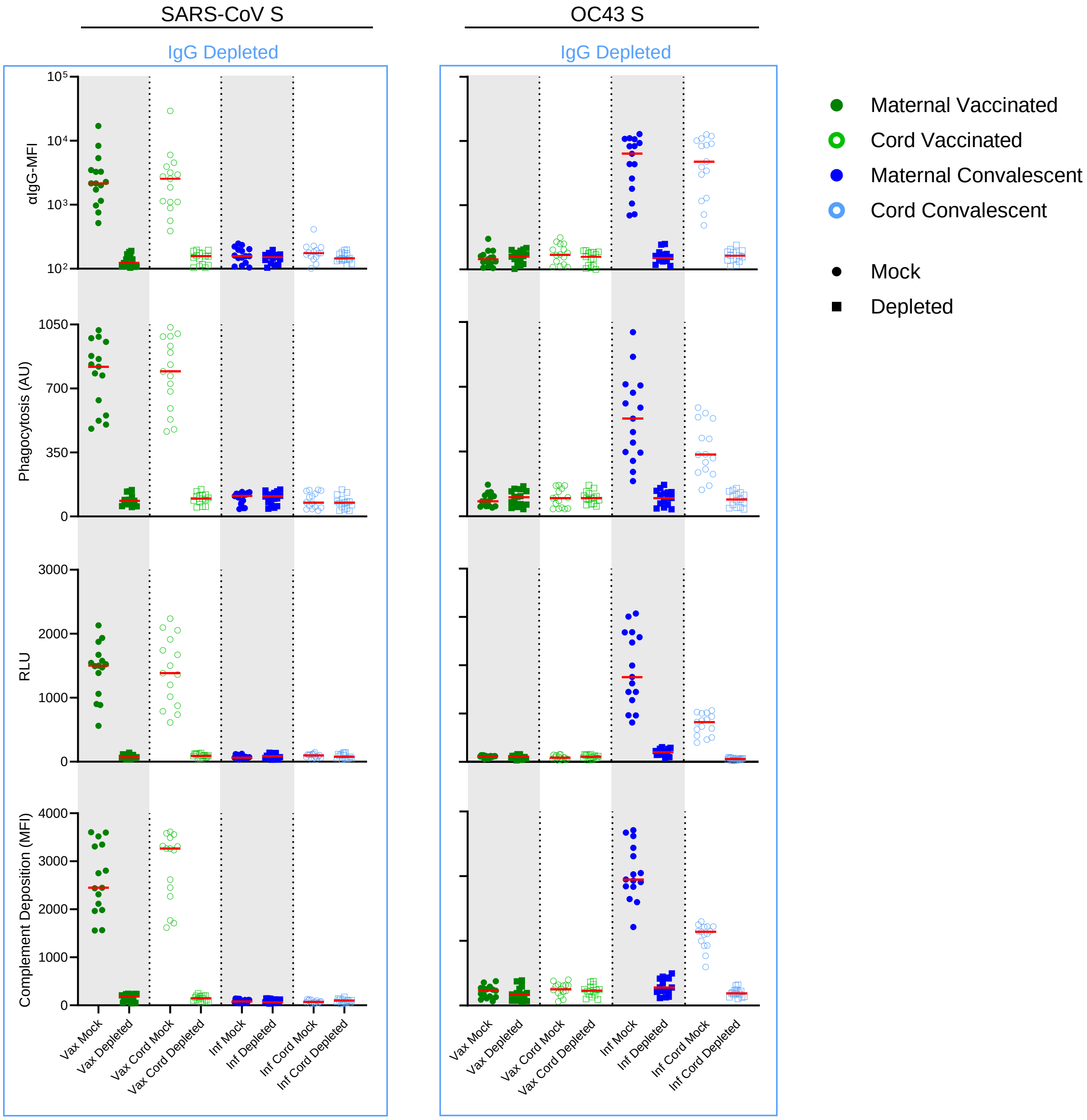

**Supplemental Figure 11. Effect of IgG depletion on effector function observed for SARS-CoV-1 and OC43 S**. Maternal (filled) and cord (open) samples among convalescent (n = 15, blue) or vaccinated (n = 15, green) individuals were depleted of IgG and tested for IgG binding against SARS-CoV-1 S (left) and OC43 S (right). ADCP, ADCC, and ADCD effector functions were measured on depleted and mock control samples. Bar indicates median.

**Supplemental Table 1. Cohort characteristics**. Dash indicates not applicable or not available, and IQR indicates interquartile range.

| Characteristic | Pregnant Convalescent<br>n=38 | Pregnant Vaccinated<br>n=50 | Naïve controls<br>n=37 |
|---|---|---|---|
| Median age (IQR), years | 31 (27-35) | 32 (29-35) | 39 (28-50) |
| Sex (n, %) | | | |
| Female | 38 (100%) | 50 (100%) | 22 (58%) |
| Male | 0 (0%) | 0 (0%) | 16 (42%) |
| Median days since PCR+ or symptom onset (IQR) | 49 (22-78) | - | - |
| Median days since second vaccine dose (IQR) | - | 20 (12-29) | - |
| Location | Belgium | Israel | US |
| IRB | CHU St. Pierre | Hadassah Medical Center | BioIVT clinical sites |
| Collection period | June 2020 – December 2020 | February 2021 | October 2020 |
| Symptoms or positive test | March 2020 – November 2020 | - | - |
| Predominant strain | Wuhan | - | - |
| Days from 2$^{nd}$ vaccine dose to delivery (cord blood sample collection) (IQR) | - | 40.5 (32-50) | - |
| Days from infection to delivery (cord blood sample collection) (IQR) | 42 (27-78) | - | - |

**Supplemental Table 2. Fc Array reagents and test conditions**

| Antigen | Source | Fc Detection | Source | Serum dilution(s) |
|---|---|---|---|---|
| SARS-CoV-2 S | Acro Biosystems SPN-C82E9 | a- IgG | Southern Biotech 2048-09 | (1:2500) 1:5000 |
| SARS CoV-2 RBD | BEI Resources NR-52366 | a-IgG1 | Southern Biotech 9054-09 | 1:1000 |
| SARS-CoV-2 S Alpha (B.1.1.7) | Sino Biological 40589-V08B6 | a-IgG2 | Southern Biotech 9070-09 | 1:250 |
| SARS-CoV-2 S Beta (B.1.351) | Sino Biological 40589-V08B7 | a-IgG3 | Southern Biotech 9210-09 | 1:250 |
| SARS-CoV-2 S Gamma (P.1) | Sino Biological 40589-V08B8 | a-IgG4 | Southern Biotech 9200-09 | 1:250 |
| SARS-CoV-2 S Delta (B.1.617.2) | Sino Biological 40589-V08B12 | a-IgA | Southern Biotech 2050-09 | 1:250 |
| SARS-CoV-2 S Omicron (B.1.1.529) | Sino Biological 40589-V08H26 | a-IgM | Southern Biotech 9020-09 | 1:250 |
| SARS-CoV-2 RBD Alpha (B.1.1.7) | Sino Biological 40592-V08H82 | FcγR2aR131 | Boesch, et. al, 2014 | 1:5000 |
| SARS-CoV-2 RBD Beta (B.1.351) | Sino Biological 40592-V08H4 | FcγR2b | Boesch, et al., 2014 | 1:5000 |
| SARS-CoV-2 RBD Gamma (P.1) | Sino Biological 40592-V08H86 | FcγR3aV158 | Boesch, et al., 2014 | 1:5000 |
| SARS-CoV-2 RBD Delta (B.1.617.2) | Sino Biological 40592-V49H-B | FcγR3bNA2 | Boesch, et al., 2014 | 1:5000 |
| SARS-CoV-2 RBD Omicron (B.1.1.529) | Sino Biological 40592-V08H121 | FcαR | Butler et al., 2021 | 1:250 |
| SARS-CoV S | Sino Biological 40634-V08B | C1q | Sigma Aldrich C1740 | 1:250 |

| | |
|---|---|
| MERS S | Sino Biological 40069-V08B-B |
| OC43 S | Sino Biological 40607-V08B |
| OC43 S-2P | Butler et al., 2021 |
| OC43 S2 | Sino Biological 40069-V08B |
| 229E S | Sino Biological 40605-V08B |
| 229E S1 | Sino Biological 40601-V08H |
| HKU1 S | Sino Biological 40606-V08B |
| HKU1 S1 | Sino Biological 40602-V08H |
| NL63 S | Sino Biological 40606-V08B |
| NL63 S1 | Sino Biological 40604-V08H |
